# A flexible agent-based modelling framework of multi-serotype pneumococcal carriage to evaluate vaccine strategies in large populations

**DOI:** 10.1101/2025.05.22.25327965

**Authors:** Nefel Tellioglu, David J. Price, Xinghui Chen, Violeta Spirkoska, Yingying Wang, Robert Moss, Natalie Carvalho, Kylie S. Carville, Patricia T. Campbell, Jodie McVernon

## Abstract

*Streptococcus pneumoniae* is a multi-serotype pathogen causing a substantial global burden of pneumonia and invasive bacterial disease. Pneumococcal conjugate vaccines have been implemented worldwide, protecting against colonisation and disease from selected high-burden serotypes, but in many settings short-term effectiveness has been eroded by serotype replacement. Vaccines with broader serotype coverage have been developed, at the cost of immunogenicity against single serotypes. To determine the longer-term impact of these new formulations, we propose a novel modelling framework that evaluates the relationship between serotype-specific vaccine immunogenicity and effectiveness, comprising immunity, carriage and clinical models. Dynamics of individual serotypes are modelled over time, advancing existing modelling paradigms that group all serotypes according to vaccine type. This framework is sufficiently flexible to consider diverse populations, emerging vaccine formulations and implementation strategies. A retrospective analysis of sequential introduction of 7- and 13-valent pneumococcal conjugate vaccines in Australia demonstrates the flexibility of the model.

## 1. Introduction

*Streptococcus pneumoniae (S. pneumoniae*) colonises the nasopharynx, but can also cause infection with both non-invasive disease, such as otitis media and pneumonia, and invasive pneumococcal disease (IPD), including meningitis and bacteraemia [1,2].

Pathogenicity is largely, though not solely, determined by the polysaccharide capsular antigen (serotype) [1,2]. Over 100 pneumococcal serotypes have been identified, with varying prevalence by age and geographic location [2,3]. Immunogenicity characteristics, such as immunoglobulin G (IgG) concentration required for protection and potential to cause invasive disease, also vary by serotype [2,4,5]. The substantial global burden of morbidity and mortality caused by pneumococcus is predominantly borne in low- and middle-income countries, and disproportionately affects children [6,7]. Older adults and children under 5 are also at risk in high-income settings [8,9].

The notable burden of disease has driven development of several pneumococcal vaccines over time, administered according to different vaccine schedules [3,5,10]. The first vaccines to be developed comprising polysaccharide capsular antigens were poorly immunogenic in children aged <2 years [11,12]. Moreover, pneumococcal polysaccharide vaccines (PPVs) only activate the humoral arm of the immune system and fail to induce immune memory [10,13]. Consequently, while PPVs provide protection against IPD, they show limited and variable efficacy in preventing community acquired pneumonia (CAP) and do not reduce nasopharyngeal colonisation, thereby failing to interrupt transmission [14]. These limitations were overcome by the development of pneumococcal conjugate vaccines (PCVs), in which polysaccharides are conjugated to a carrier protein [10,13].

Conjugation enhances immunogenicity and effectiveness by inducing immune memory, including in young children [10]. Conjugate vaccines protect against both systemic and mucosal infection and prevent nasopharyngeal colonisation by vaccine serotypes, thereby reducing transmission in the community [15,16]. A seven valent conjugate vaccine (7vPCV), first licensed in the USA in 2000, included seven serotypes causing at least 70% of disease in young children in high-income countries [2,15]. However, serotype replacement and continuing burden of disease has driven the creation of higher valency vaccines [7,10,15,16]. A 13vPCV became available in 2010, and more recently 15vPCV, 20vPCV, and 21vPCV have been developed [4,10,15]. However, there is some concern regarding declining immunogenicity against shared serotypes with higher-valency PCVs, with uncertain implications for effectiveness [15,17].

Variability in serotype-specific immunogenic response of multiple vaccine products necessitates a flexible approach to modelling potential impacts of different vaccines. While previous models examining use of pneumococcal vaccines account for the multi-serotype nature of *S. pneumoniae* to some degree, they often depend on simplifying assumptions about serotypes (vaccine-type groupings, or inclusion of a limited number of serotypes), co-infections, immune response, and host representation (only considering a specific age group) [18–29]. Models that group serotypes into vaccine and non-vaccine types typically assume that there would be no co-infection with serotypes from the same group, which does not allow co-infection with two non-vaccine serotypes (e.g., [19,28,29]). This assumption has the potential to alter transmission rates once vaccine-type serotype circulation decreases following vaccine introduction. Additionally, these models often assume a homogeneous vaccine-induced immune response to vaccine-type serotypes (e.g., [18,23,24]), which does not allow observed differences in serotype-specific immunogenicity to be incorporated. These assumptions and simplifications make it challenging to fully assess the effectiveness of vaccine programs across groups receiving different vaccines, as well as the potential implications of reduced immunogenicity of higher valency vaccines on pneumococcal disease outcomes.

There is a need to realistically capture serotype- and age-specific heterogeneities in transmissibility and vaccine-induced immunogenicity to estimate effectiveness of vaccine policies at the population level. To achieve this, we have developed a flexible individual-level modelling framework that can incorporate individual serotype transmission, serotype-specific vaccine immunogenicity, and any combination of vaccines targeting different serotypes in various age groups. Using this model, with parameters matched to historical observations, we show that 13vPCV introduction averted further disease outcomes compared to the hypothetical scenario of continued 7vPCV use in Australia. The framework is readily adaptable to different settings and can be used to simulate future scenarios to determine the likely impact of changes to pneumococcal schedules, products and target populations.

## 2. Methods

Our modelling framework is comprised of three integrated models (Fig 1):

1. **An immunity model**: of immune responses within individuals, incorporating available evidence on serotype- and age-specific initial levels of vaccine-induced antibody titres and waning dynamics of immune responses,
2. **A transmission model**: capturing transmission amongst the population of multiple serotypes of pneumococcal infection (carriage),
3. **A clinical model**: representing the progression of infection to clinical outcomes of importance, namely IPD and CAP, according to serotype. The clinical model incorporates available evidence on serotype- and age-specific IPD and age-specific CAP outcome rates.

**Fig 1:**
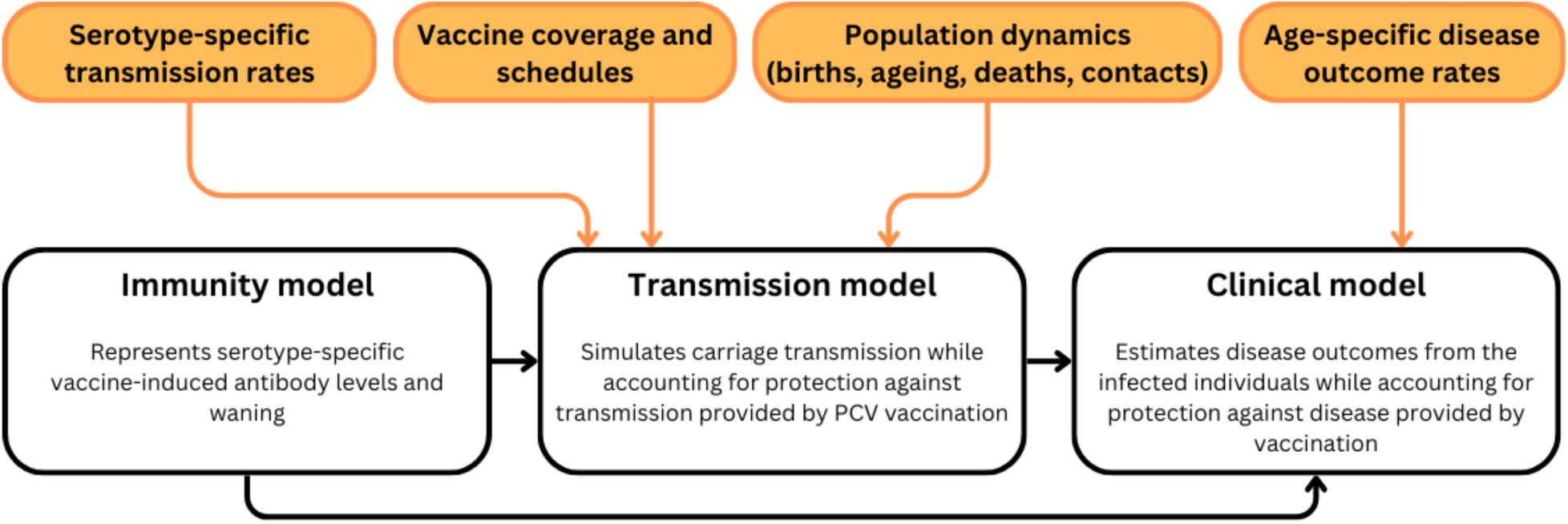
Modelling procedure diagram. In the immunity model, an individual’s vaccine-induced antibody responses are calculated based on the vaccine product received, number of doses administered, and time since vaccination. Antibody levels post-vaccination and antibody decay over time are informed by evidence from literature (see Section 2.1.1 and Supplementary Material section S1). The probabilities of infection acquisition and disease development are then calibrated to historical data. In the transmission model, age-specific carriage of individual serotypes is simulated, where individuals can carry up to two distinct serotypes. The clinical model takes infected individuals from the transmission model, and the disease outcomes are simulated based on individuals’ vaccine status, age, and the serotype they carry.

In the following, we provide a detailed explanation of the immunity, transmission, and clinical models, the model calibration and the scenarios we consider for our subsequent analyses.

### 2.1. Immunity model

An individual’s initial vaccine-induced antibody response to each serotype is drawn from a distribution derived from literature (see Section 2.1.1), and these antibodies wane over time (described below). At the time of exposure to an infected individual (determined within the transmission model), the individual’s antibody level modifies their probability of being infected. If they become infected, their antibody level further informs their probability of progressing to disease (within the clinical model). Following existing evidence in the literature, we assume that probabilities of acquisition and developing disease follow a logistic function [30].

Fig 2 below provides an example of this process for a single individual.

**Fig 2:**
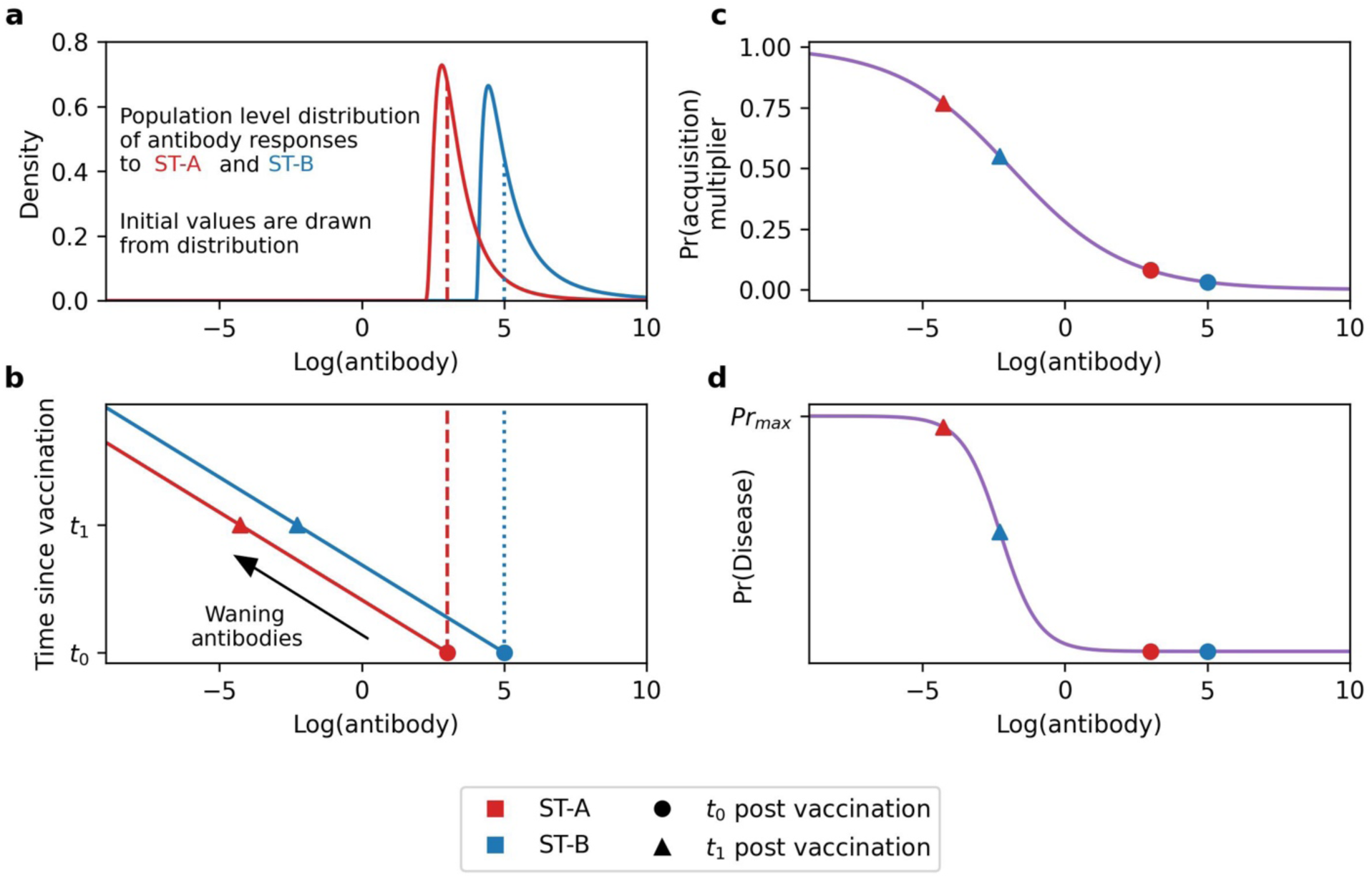
A schematic of the vaccine-induced antibody levels for a given vaccine product and two exemplar serotypes (ST-A and ST-B) in the immunity model. In this example, we assume that ST-A and ST-B are both vaccine-type serotypes. Panel a shows the population level distribution of initial antibody responses to ST-A (red) and ST-B (blue).

Dashed and dotted lines are the sampled initial antibody responses to ST-A (red) and ST-B (blue), respectively, for a given individual, shown as filled circles in panel b. Panel b shows the calculation of waned log-antibody titres from t_0_ to t_1_ post-vaccination. Panel c shows the probability of acquisition multiplier given log-antibody titres, which is used in the transmission model. Panel d shows the probability of developing disease (IPD or CAP) given infection, depending on log-antibody levels to the infecting serotype (ST-A or ST-B).

We assume that in individuals exposed to serotype’*a*’, for which they have no immunity, the probability of acquisition multiplier, 𝑉_!_, is 1. If they have vaccine-induced immunity against serotype *a*, which can be only provided by conjugate vaccines, their probability of acquisition multiplier is calculated using a logistic function:

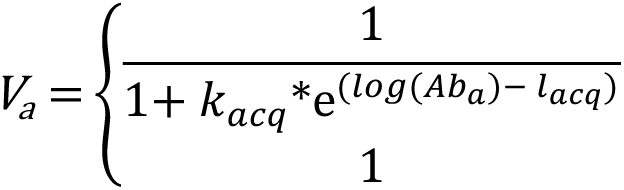 if vaccinated with conjugate vaccine containing serotype *a*, otherwise

where 𝑙𝑜𝑔(𝐴𝑏_a_) is log-antibody against serotype *a*, 𝑘_*acq*_ and 𝑙_*acq*_ are the logistic curve’s growth rate parameter and sigmoid midpoint, respectively.

As above, we assume that individuals have a baseline probability of developing disease that is modified by their antibody response according to the following equation for the probability of developing disease multiplier, 𝑉_*a,dis*_:

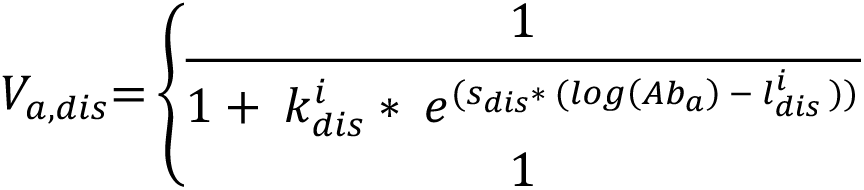 if vaccinated with vaccine containing serotype *a* otherwise

where 𝑠_dis_ is a scaling parameter and 𝑘^i^_dis_ and 𝑙_dis_^i^ are the logistic curves growth rate parameter and sigmoid midpoint for age group *i*, respectively. We assume that 23vPPV does not provide any protection against developing CAP.

#### 2.1.1 Vaccine-induced Immunity Model

We conducted a rapid review of studies reporting *S. pneumoniae* antibody levels post-vaccination that revealed considerable differences in immune responses by serotype (Supplementary Material Section 1.1). Briefly, we recorded antibody geometric mean concentrations (GMCs) with 95% confidence intervals for IgG from the eligible studies. Unlike previous models that assume the same protection against serotypes included in the same vaccine, we introduce a biologically-informed grouping of serotypes into low, moderate, and high immunity categories based on this review. Serotypes were categorised based on the magnitude of the antibody responses, for each vaccine product and dose combination across multiple studies, and an estimate of the antibody response was calculated for each grouping. Post-vaccination IgG antibody responses to Serotype 3 were consistently lower than other serotypes, necessitating a separate category [15,31]. Full details of the rapid review are provided in Supplementary Material Section 1.1.

Given inconsistent and limited information on cross-reactivity of protection against serotypes that are not included in vaccine products, we assume that antibodies are generated only against serotypes in the vaccine product.

#### 2.1.2 Naturally-acquired Immunity

Given the absence of evidence for subsequent protection against infection and disease, many pneumococcal modelling studies do not consider natural immunity following infection [19,21,32]. Similarly, our model does not include a boost to antibodies following infection, with individuals returning to the susceptible state following clearance of infection with no naturally-acquired protection. We do, however, indirectly account for the accumulation of protection against pneumococcus over the life course by incorporating the observed reduction of carriage duration throughout late childhood and adult life (Supplementary Material Section 4.2).

### 2.2. Transmission model

The transmission model is an individual-based model that incorporates demographic and transmission dynamics. Unlike earlier pneumococcal transmission models that aggregate serotypes based on vaccine type [19,21,28,29], our framework simulates transmission of individual *S. pneumoniae* serotypes. This approach allows for realistic modelling of co-infections with multiple vaccine serotypes, multiple non-vaccine serotypes or a mix of both, not possible in transmission models that group serotypes. As noted in Section 2.1, we assume a susceptible-infectious-susceptible (SIS) model for *S. pneumoniae* infection, where “susceptible” represents individuals not carrying *S. pneumoniae*, and “infectious” represents individuals carrying at least one serotype of *S. pneumoniae*. Disease outcomes are considered separately in the clinical model. Similar to previous modelling studies, the infectious duration for an individual is sampled from an exponential distribution with an age-specific mean infectious duration (Table 1) [18,21,29,33]. Upon recovery, infectious individuals re-enter the susceptible class, with protection derived from antibodies according to the time since their last vaccination (as dictated by the immunity model).

**Table 1:** Fixed parameters.

| Parameter | Description | Values | Notes/<br>Reference |
| --- | --- | --- | --- |
| Age-specific duration of infection (days) | Exponentially distributed with age-specific mean | 0–2y: 72,<br>2–4y: 28,<br>5–17y: 18,<br>18+y: 17 | [18,21,29,33] |
| Age specific probability of developing IPD when developing disease | This probability value is used to decide whether someone will develop IPD when they develop disease (IPD or CAP). | 0-1m:0.314,<br>2-3m: 0.341,<br>4-5m: 0.368,<br>6-11m:0.279,<br>1-4y: 0.237,<br>5-14y: 0.173,<br>15-24y: 0.127,<br>25-49y: 0.136,<br>50-64y: 0.117,<br>65-74y: 0.073,<br>75-79y: 0.051,<br>80+y: 0.041 | Values are estimated by age-specific IPD and CAP incidence values between 2006–2018 provided by NCIRS and population size taken from Australian Bureau of Statistics (ABS) [34] |
| Age-specific waning half-life of antibody levels (days) | - | 0-17-y: 125<br>18+-y: 600 | Supplementary Material Section 1 |
| External exposure | For X times a year, Y% of the population is exposed to Z randomly selected serotypes. Individuals become infected based on the antibody levels. | X: 4,<br>Y: 0.06,<br>Z: 2 | The values are set so that the baseline prevalence of serotypes without any vaccine introduction stays at an endemic level. |
| Reduction in susceptibility to co-infections (C) | Fractional reduction in susceptibility (between 0–1) | 0.8 given one infection,<br>1 given two infections (maximum number of co-infections is two) | [26] |
| Initial number of circulating serotypes | 7vPCV, 13vPCV and 23vPPV serotypes are assumed to be circulating in the population, along with some non-vaccine serotypes. | 55 | The value is set to observe at least the number of unique serotypes in Dunne et al. (2016) [40] and number of unique serotypes in IPD cases |
| Percentage of vaccine type serotypes carriage among all carriages at model initialisation | - | 7vPCV: 55%,<br>13vPCV/non 7vPCV: 25%,<br>23vPPV/non 13vPCV: 15%,<br>Other serotypes (NVT): 5% | [40] |
| Age-specific initial fraction of infected population | - | 0–2y: 24%,<br>2+y: 12% | [40] |

#### 2.2.1 Demography

The model includes births, ageing, age-specific mortality, and age-specific migration to capture the age distribution of a population over time (Supplementary Material Section 2.1). It takes inputs of initial population size, initial age distribution, age-specific death rates, annual net birth rates, annual migration rates, and age distribution in the emigrated population to simulate demographic dynamics. We sourced the inputs from Australian Bureau of Statistics and Macro Trends Global to align with Australian population dynamics (Supplementary Material Section 4.1) [34,35].

#### 2.2.2 Mixing assumptions

Transmission within and between age groups is driven by contact rates, derived from synthetic contact matrices (Supplementary Material Section 2.2.1). In this work, we have assumed contacts in our population according to the contact matrices provided by Prem et al. [36].

#### 2.2.3 Carriage assumptions

Carriage of serotypes is determined in multiple steps. We first determine if an individual will be exposed to any serotype (step 1), we next determine the serotype (step 2), then assess the individual’s serotype-specific vaccine-induced antibody levels (step 3) and current infections (step 4), to determine if the individual gets infected by a serotype (step 5).

In step 1, we estimate the probability of exposure to any of the currently circulating serotypes for each individual. In this study, we define “exposure” as an individual coming into contact with a serotype. This exposure may lead to an infection depending on other factors (i.e., individual’s immunity, existing co-infections). Individual infection risk in a time step is determined by serotype prevalence, age, contact rates, current infection status and current antibody levels:

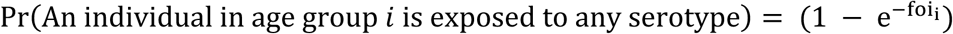

where, foi_5_ represents the force of infection on an individual in age class *i*.

The force of infection on an individual in age group *i* comprises age-specific community transmission:

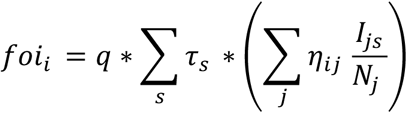

where:

- 𝑞 = Transmission coefficient
- 𝜏_s_ = Transmission coefficient multiplier of serotype *s*
- 𝜂_ij_ = Number of daily community contacts between an individual in age group *i* and individuals in age group *j*
- 𝐼_js_ = Number of individuals in age group *j* infected with serotype *s*
- 𝑁_j_ = Number of individuals in age group *j*

In step 2, we assume that an individual can be only newly infected by a single serotype at every time step. The exposed serotype, serotype *a,* is randomly selected from the distribution of prevalences of individual serotypes in the population weighted by their transmission coefficient multipliers. In steps 3 and 4, we then determine whether the individual is going to be infected by *S. pneumoniae* serotype *a* by checking the individual’s vaccine-induced antibody levels and existing infections:

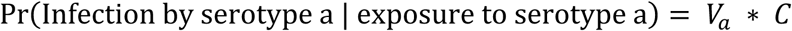

where 𝑉_a_ and 𝐶 are the vaccine-induced probability of acquisition multiplier (between [0,1]) to serotype *a* (see Section 2.1) and the reduction of susceptibility given existing infections (between [0,1]), respectively.

We allow co-infections in the model assuming that individuals may be infected with multiple serotypes concurrently, with a reduced probability of acquiring another serotype if already infected, representing within-host competition. In this work, we assumed that 𝐶 takes a value of 0 if the individual is already infected by the exposed serotype. We also assumed that 𝐶 takes values of 1 (fully susceptible), 0.8 and 0 for the existing number of infections of 0, 1, and 2, respectively, which means that individuals could be infected with a maximum of two serotypes concurrently (Supplementary Material Section 2.2.4).

In addition to community-level circulation, we introduce serotypes externally via two processes. First, an external exposure rate simulates the ongoing introduction of serotypes through population migration, assumed to have the same age-specific and serotype-specific incidence of *S. pneumoniae* carriage as the local population to maintain the same level of serotype- and age-specific carriage while population size increases.

Second, we introduce serotypes sampled uniformly from the set of all possible serotypes which may not be currently circulating within the local population, which allows possibility of serotype replacement under immune selection over time.

This approach enables us to simulate transmission of individual serotypes and vary serotype-specific transmission rates to match the historically observed dominance of specific serotypes.

#### 2.2.4 Vaccination rollout programs

The model simulates the implementation of historical vaccine schedules with their rollout years, eligible age range, annual fraction of on-time and late vaccine coverages of the eligible age group, number of doses and time interval between doses. We assumed that individuals receiving the first dose on time would also receive the following doses on time. We also assumed that maximum protection provided by a vaccine starts immediately. For the simulation study, we consider vaccinations schedules shown in Table S8.

### 2.3. Clinical model

The clinical model takes information on the age and vaccine history of currently infected individuals from the transmission model, and the antibody levels at the current time, derived from these values using the immunity model. The probability of developing disease is of the form:

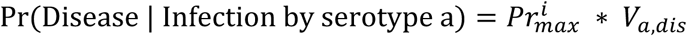

where 𝑃𝑟_*max*_^*i*^ and 𝑉_*a,dis*_ are the maximum probability of developing disease given infection for age group *i* and the probability of developing disease multiplier for individuals infected by serotype *a* (described in Section 2.1), respectively.

In this work, we assumed that 23vPPV does not provide any protection against developing CAP [14], therefore, we separate individuals vaccinated with 23vPPV in the clinical model (see Fig S4 in Supplementary Material). We assume that an individual can develop disease only once from a given infection with a single serotype. Therefore, at each time step, we consider only individuals who have not already developed disease during their current infection period. We check if the infected individual has vaccine-induced antibody levels against their infecting serotypes. Then, by using the probability of developing disease formulation, the individual’s vaccine-induced antibody levels and age (see Section 2.1), the model determines whether an infected individual develops a disease outcome. The clinical model splits these disease outcomes into IPD and CAP based on the relative probability of each occurring. In this work, the age-specific IPD to CAP ratio is calculated from the age-specific IPD and CAP incidences provided by the National Centre for Immunisation Research and Surveillance (NCIRS). The specific disease outcome, when it occurred and other demographic details are recorded in the model (Supplementary Material Section 3) [14].

### 2.4. An analysis of 13vPCV in Australia, 2002–2018

As a simulation study, we aimed to estimate the impact of the 13vPCV rollout in comparison to the continuation of 7vPCV after 2011. We compared the disease burden (IPD and CAP) across the total population in two scenarios: “With 13vPCV” (Australian vaccination strategy, 7vPCV between 2005–2010 and 13vPCV between 2011–2018) and “Without 13vPCV” (7vPCV between 2005 and 2018) in the non-Indigenous Australian population.

#### 2.4.1 Data

We used historical vaccine coverage and disease rates in the non-Indigenous Australian population (Supplementary Material Section 4.2). We used historical vaccine coverage from Kabir et al (2021), Frank et al (2020) and NCIRS data (Supplementary Material 4.2.2) and disease rates from NCIRS (Section 3.1) [37,38]. To calculate disease rates, we used serotype- and age-specific IPD cases. In Australia, data for historical *S. pneumoniae*-specific CAP cases were not available and laboratory testing is often not performed; we approximated this by pooling data for International Classification of Diseases (ICD) codes J13 (Pneumonia due to S. pneumoniae), J18.1 (Lobar pneumonia) and 11% of pneumonia due to an unspecified cause (J15.9 and J18 except J18.1) [39].

#### 2.4.2 Model calibration

Modelled disease outcomes were compared with observed age-specific IPD and CAP rates and serotype proportions in IPD between 2006–2018 provided by NCIRS. The CAP data we used in this analysis rely on a fixed percentage of unspecified pneumonia, which may be influenced by changes in diagnostic practices for other diseases. Therefore, in choosing model parameters during the calibration process, we prioritised the match to age-specific IPD rates rather than matching the CAP trends due to the uncertainty in CAP data. The model was initialised in 2002 with the carriage rates of the pre-vaccination era drawn from Dunne et al. (2016), which reported the overall prevalence of carriage in children aged two years and under, as well as prevalence of carriage split into 7vPCV and 13vPCV/non 7vPCV serotypes [40]. Values of the parameters that are fixed in the analysis are provided in Table 1.

The percentage of IPD cases due to serotypes in each of 7vPCV, 13vPCV and 23vPPV; age-specific carriage prevalence of 7vPCV, 13vPCV/non 7vPCV, 23vPPV/non 13vPCV serotypes in the pre-vaccination era; and the age-specific percentage of 7vPCV, 13vPCV/non 7vPCV, 23vPPV/non 13vPCV serotypes in IPD cases between 2006–-2018 were used as summary statistics during calibration. Values of the parameters in Table 2 were calibrated to match the key trends described.

**Table 2:**
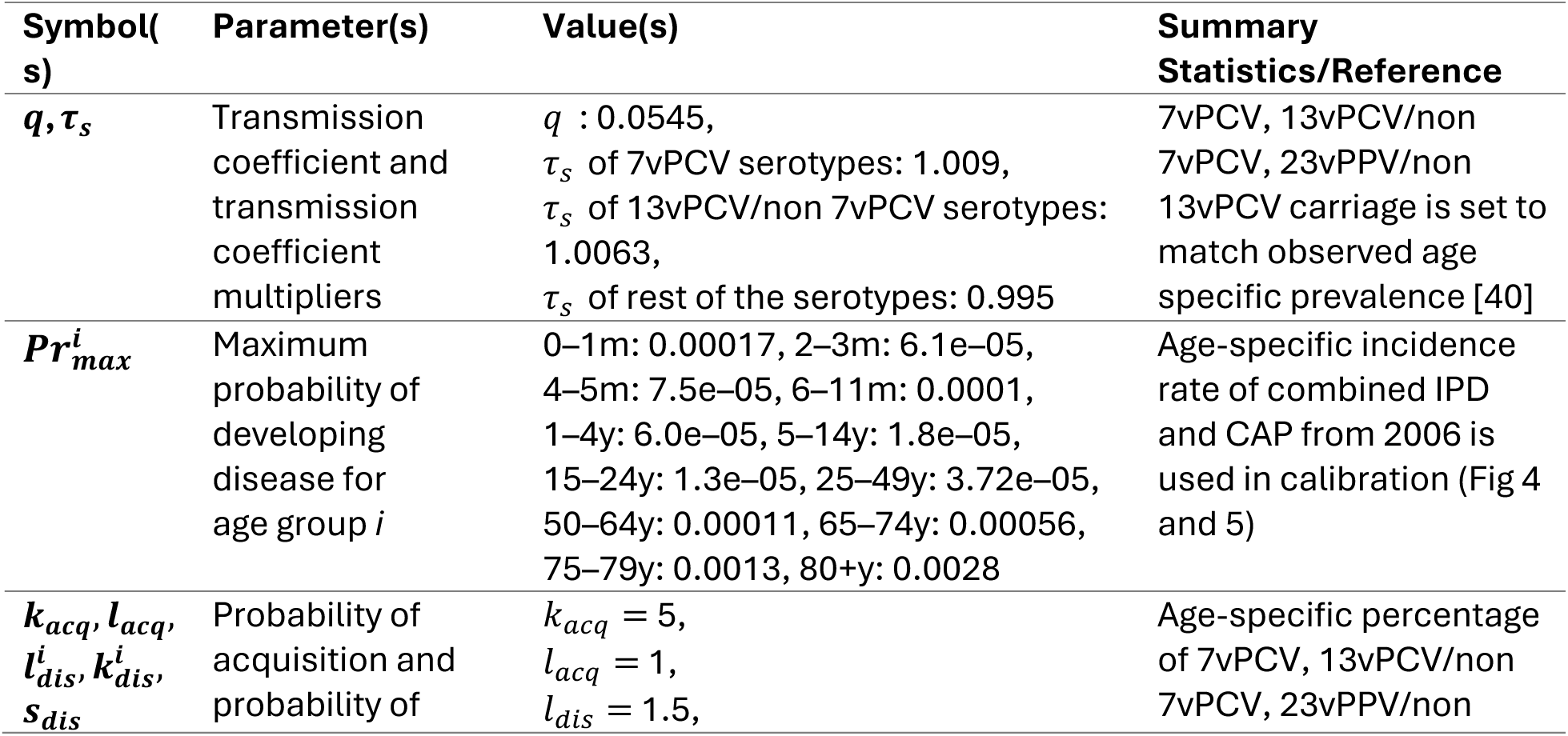
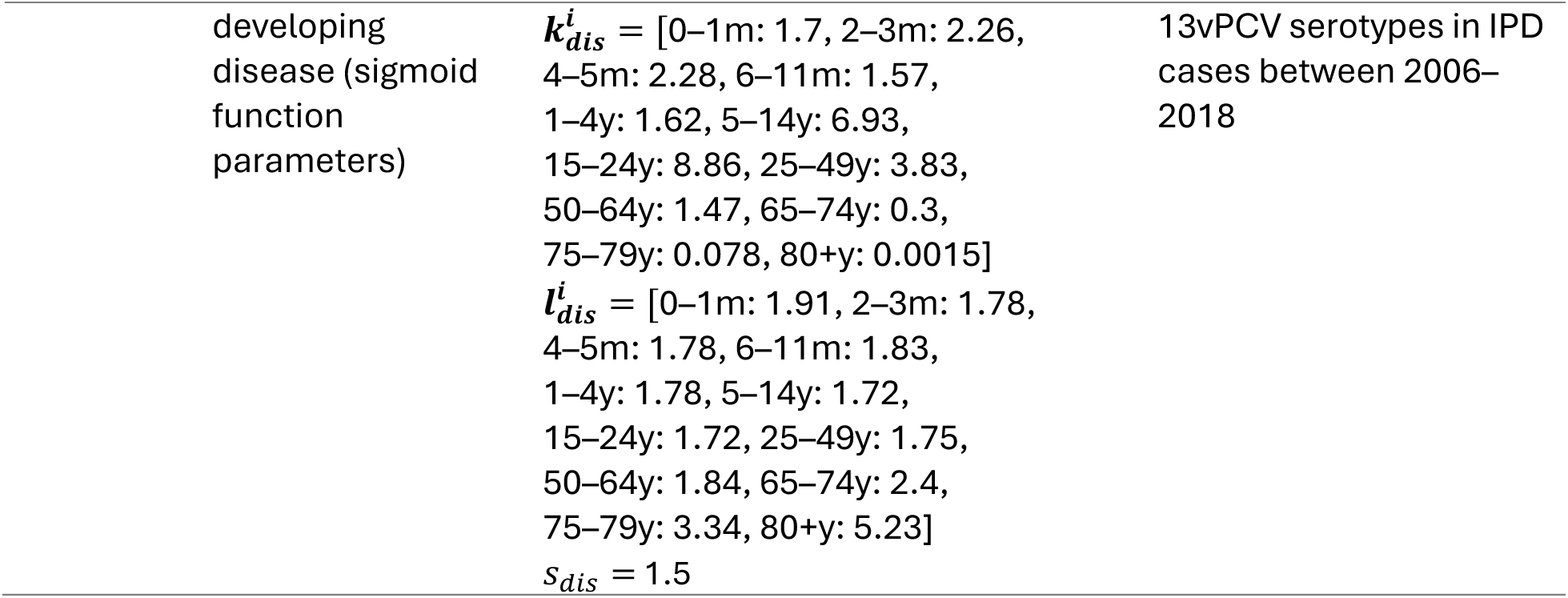
Calibrated parameters to match the observed summary statistics.

#### 2.4.3 Sequential pneumococcal vaccine introduction (7vPCV, 13vPCV) in Australia, 2002–2018

In Australia, the pneumococcal vaccine schedule was introduced in 2005 for the non-Indigenous population (paediatric: 7vPCV at 2, 4 and 6 months of age; adult: one dose of 23vPPV at age 65 or older). In 2011, the paediatric vaccine schedule was changed to 13vPCV (2, 4 and 6 months of age) with a catchup program. In the catch-up program, individuals under four years old who previously received the 7vPCV vaccine received a single dose of the 13vPCV vaccine. The 23vPPV adult vaccine rollout was maintained throughout both scenarios. The vaccine coverage data were sourced from NCIRS, Kabir et al. (2021), and Frank et al. (2020) [37,38]. In 2018, the ages at which 13vPCV was delivered changed to 2, 4, and 12 months. We limited the study period to 2002–2018 to ensure that our analysis focused solely on the effects of the product change from 7vPCV to 13vPCV.

## 3. Results

### 3.1. Model Calibration

The modelled age-stratified incidence of IPD per year, per 100,000 over the period 2002–2018 is well aligned with data (Fig 3). The age-stratified incidence of CAP per year, per 100,000 over the period 2002–2018 is shown in Fig 4. Modelled CAP results are well aligned with the data in younger age groups, but less so in later years for older age groups.

**Fig 3:**
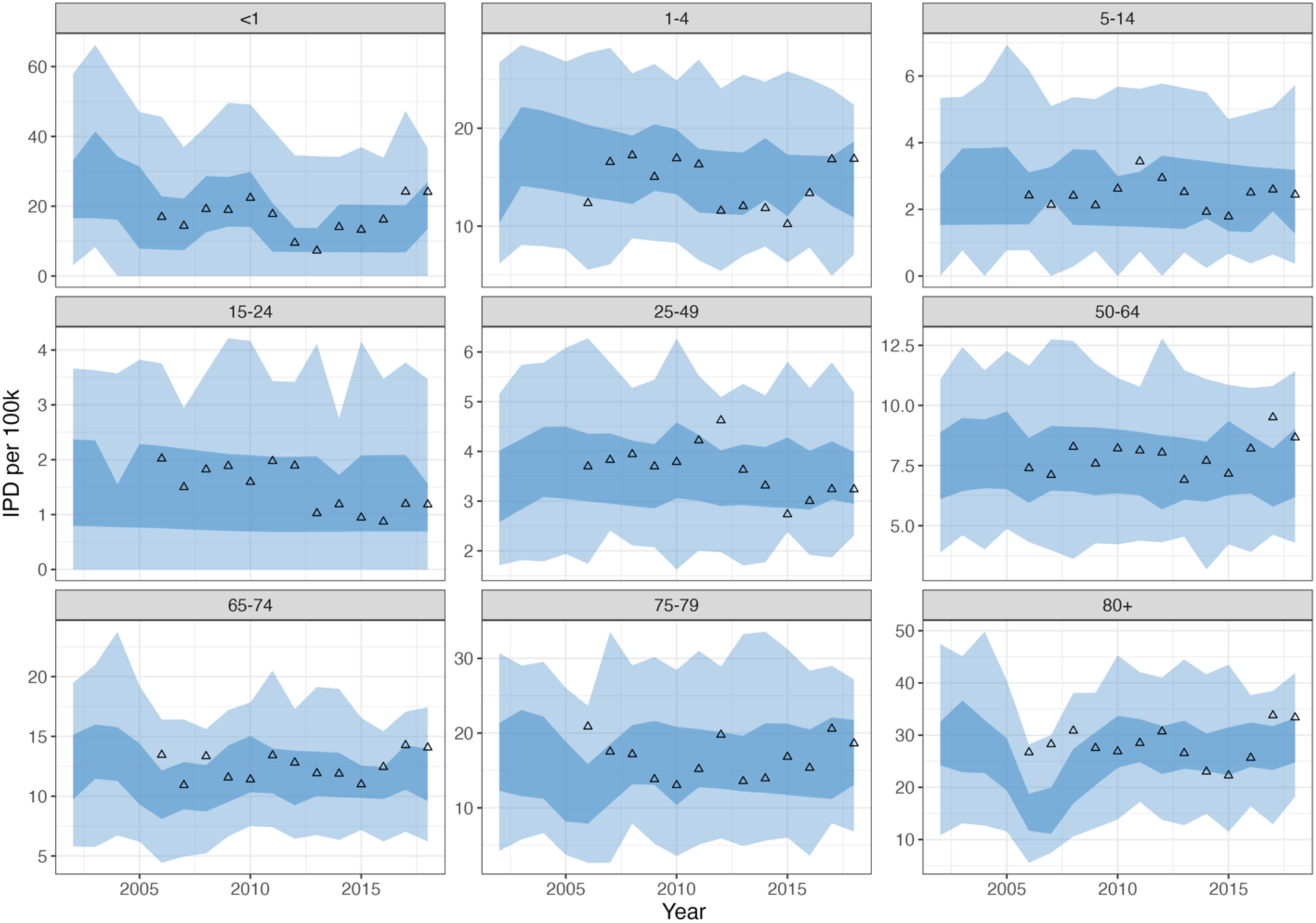
Age-stratified IPD incidence per year, per 100,000 for the period 2002–2018. The 50% (dark) and 95% (light) intervals of 100 simulation runs are shown as blue ribbons. While serotypes are modelled individually, they have been grouped for visual representation. Data from NCIRS for the non-Indigenous population are shown as triangles.

**Fig 4:**
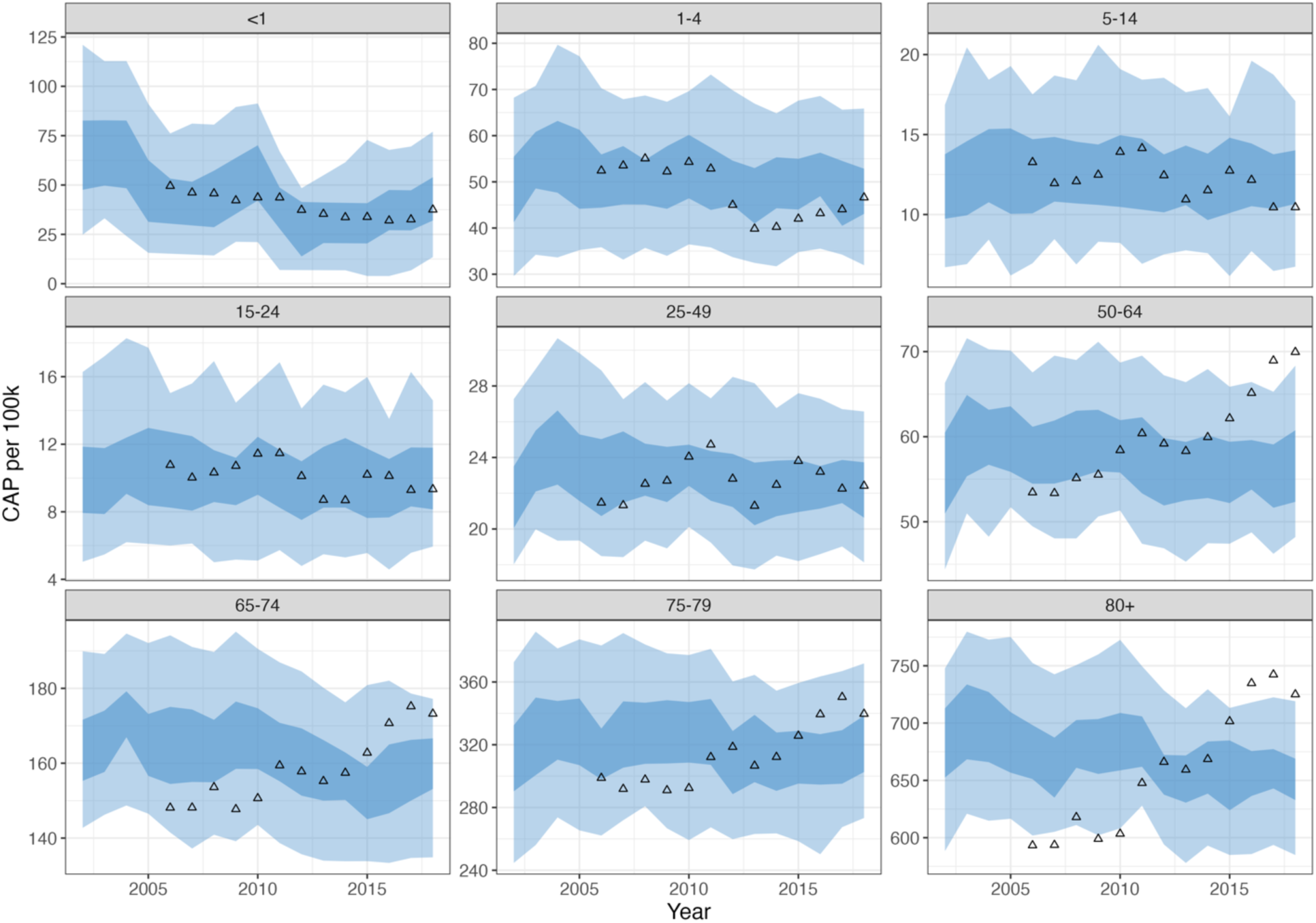
Annual age-stratified CAP incidence per 100,000 for the period 2002–2018. The 50% (dark) and 95% (light) intervals of 100 simulation runs are shown as blue ribbons. While serotypes are modelled individually, they have been grouped for visual representation. Data from NCIRS for the non-Indigenous population is shown as triangles.

However, as described in Methods 2.4.1 data for *S. pneumoniae*-specific CAP cases were not available and were approximated, including a percentage of unspecified pneumonia diagnoses. We did not aim for a perfect match to approximated data that may not be an accurate representation of true CAP cases. The proportion of vaccine-type serotypes observed in modelled IPD cases over the period 2002–2018 is well aligned with data on IPD presentations (Fig 5).

**Fig 5:**
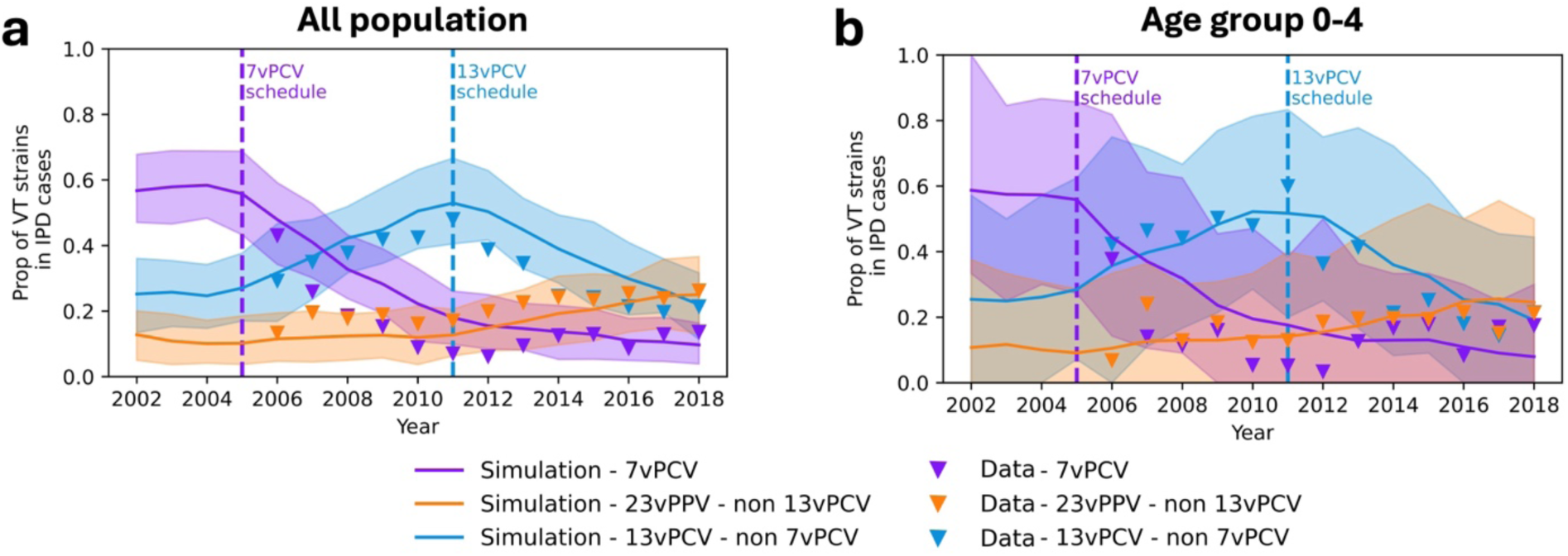
Proportion of vaccine-type serotypes in IPD cases for the period 2002–2018. Panel a reports results across all population age groups, Panel b is restricted to IPD cases in children aged 0–4 years. Both panels chart the proportion of IPD cases over time attributable to 7vPCV, 13vPCV/non 7vPCV and 23vPPV/non 13vPCV vaccine-type serotypes. While serotypes are modelled individually, they have been grouped for visual representation. 7vPCV serotypes are aggregated and shown in purple while 13vPCV/non 7vPCV and 23vPPV/non 13vPCV vaccine-type serotypes are similarly aggregated and are shown in blue and orange, respectively. Mean and 95% quantiles of 100 simulation runs are presented, and data provided by NCIRS is shown as triangles.

### 3.2. The analysis of 13vPCV in Australia, 2002–2018

Modelled carriage prevalence over the period 2002–2018 separated into 7vPCV, 13vPCV and 23vPPV serotypes captures phenomenological features of serotype replacement in pneumococcal disease known to have occurred throughout this period (Fig 6a). At the start of the funded vaccination program in 2005, modelled carriage prevalence of the serotypes in 7vPCV begins to decrease, while the non-vaccine types (NVT) at that time (i.e., 13vPCV/non 7vPCV, 23vPPV/non 13vPCV and NVT) all rise. In 2011, when 13vPCV was introduced, prevalence of included serotypes similarly decreases, while 23vPPV/non 13vPCV and NVT continue to increase. In the no 13vPCV scenario, we observe a continued increase in the carriage of 13vPCV serotypes and a decrease of 23vPPV/non 13vPCV serotypes (Fig 6b).

**Fig 6:**
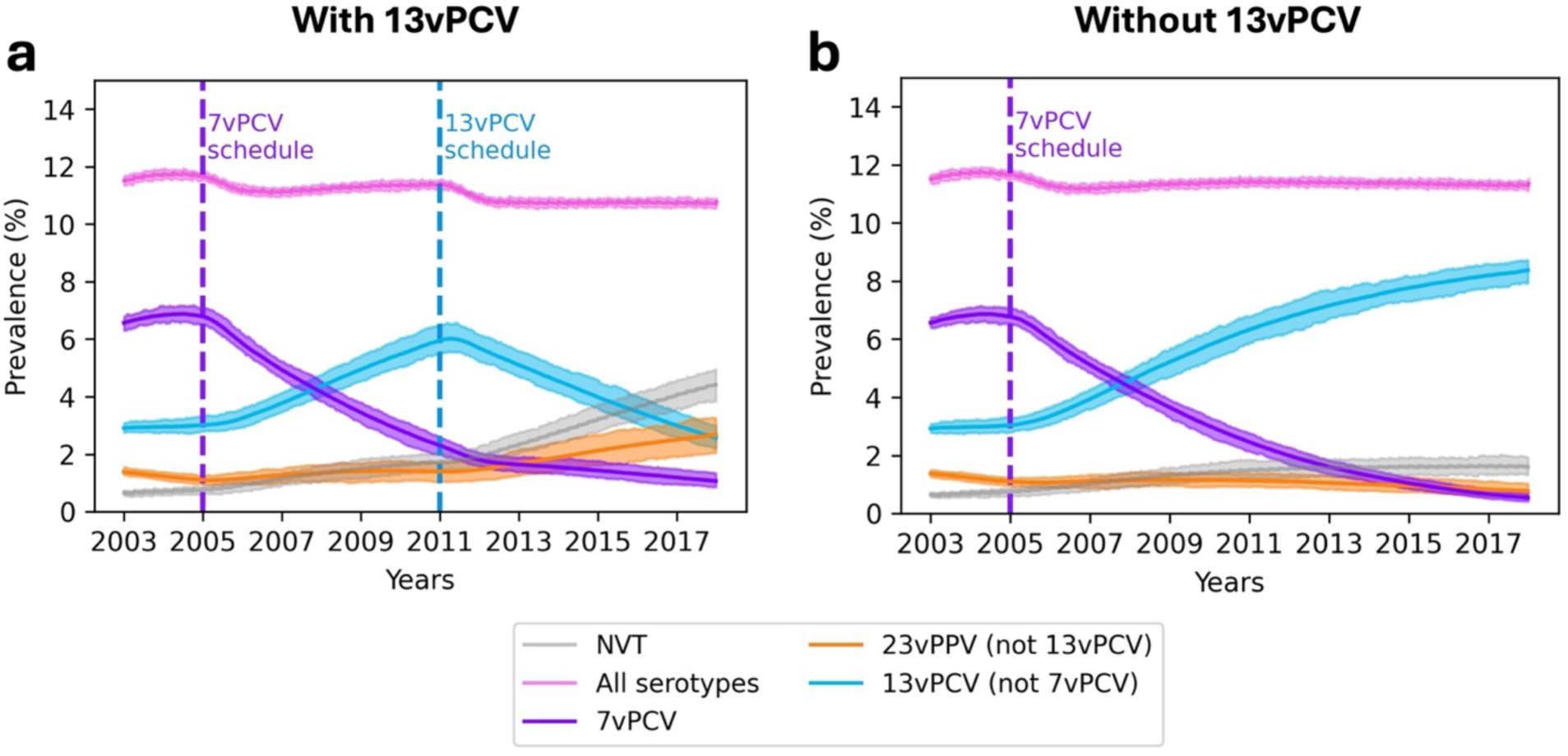
Prevalence of serotypes from 2002–2018, grouped into 7vPCV, 13vPCV/non 7vPCV, 23vPPV/non 13vPCV. While serotypes are modelled individually, they have been grouped for visual representation. Aggregated 7vPCV serotypes (purple), 13vPCV/non 7vPCV (blue), 23vPPV/non 13vPCV (orange) vaccine-type serotypes are shown alongside all aggregated (pink) and non-vaccine type (grey) serotypes. The results of 100 simulation runs are presented in each panel, with lines representing the mean for each serotype (shading represents 95% quantiles). Historical carriage data are not available in this setting as a comparison.

In the absence of 13vPCV, due to the higher transmissibility of 13vPCV serotypes compared to non-13vPCV serotypes in our model, the carriage of 23vPPV/non13vPCV serotypes decreased over time due to competition (Fig 7b). Our model suggests we would have observed a further decline in the proportion of 7vPCV serotypes in IPD cases per year, per 100,000 after 2012 (Fig 7b).

**Fig 7:**
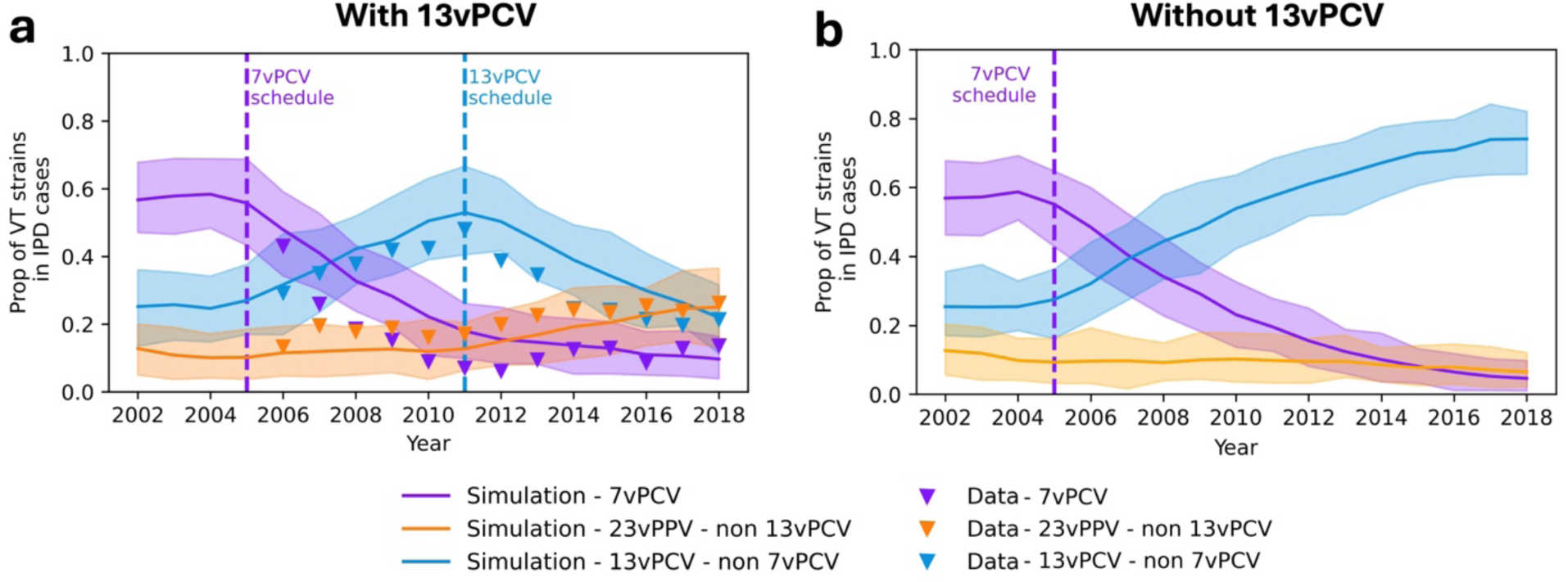
**Proportion of vaccine-type serotypes in IPD cases for the period 2002–2018 in (a) Australian vaccination strategy and (b) No 13vPCV scenario**. The results represent the whole population. While serotypes are modelled individually, they have been grouped for visual representation. 7vPCV serotypes are aggregated and shown in purple while 13vPCV/non 7vPCV and 23vPPV/non 13vPCV vaccine-type serotypes are similarly aggregated and are shown in blue and orange, respectively. Mean and 95% quantiles of 100 simulation runs are presented, data are shown as triangles.

With 13vPCV, we observed lower CAP per year, per 100,000 (79.79, 95% CI: [77.53 - 81.85]) and IPD per year, per 100,000 (6.99 95% CI: [6.38 - 7.64]) burden after 2011, respectively, compared to the no 13vPCV scenario (83.96 95% CI: [81.9 – 85.8] and 7.4 95%CI: [6.91 - 7.9], respectively; Fig 8). This analysis shows that the 13vPCV paediatric vaccine schedule in the non-Indigenous Australian population reduced the burdens of CAP and IPD by 5% and 5.5%, respectively.

**Fig 8:**
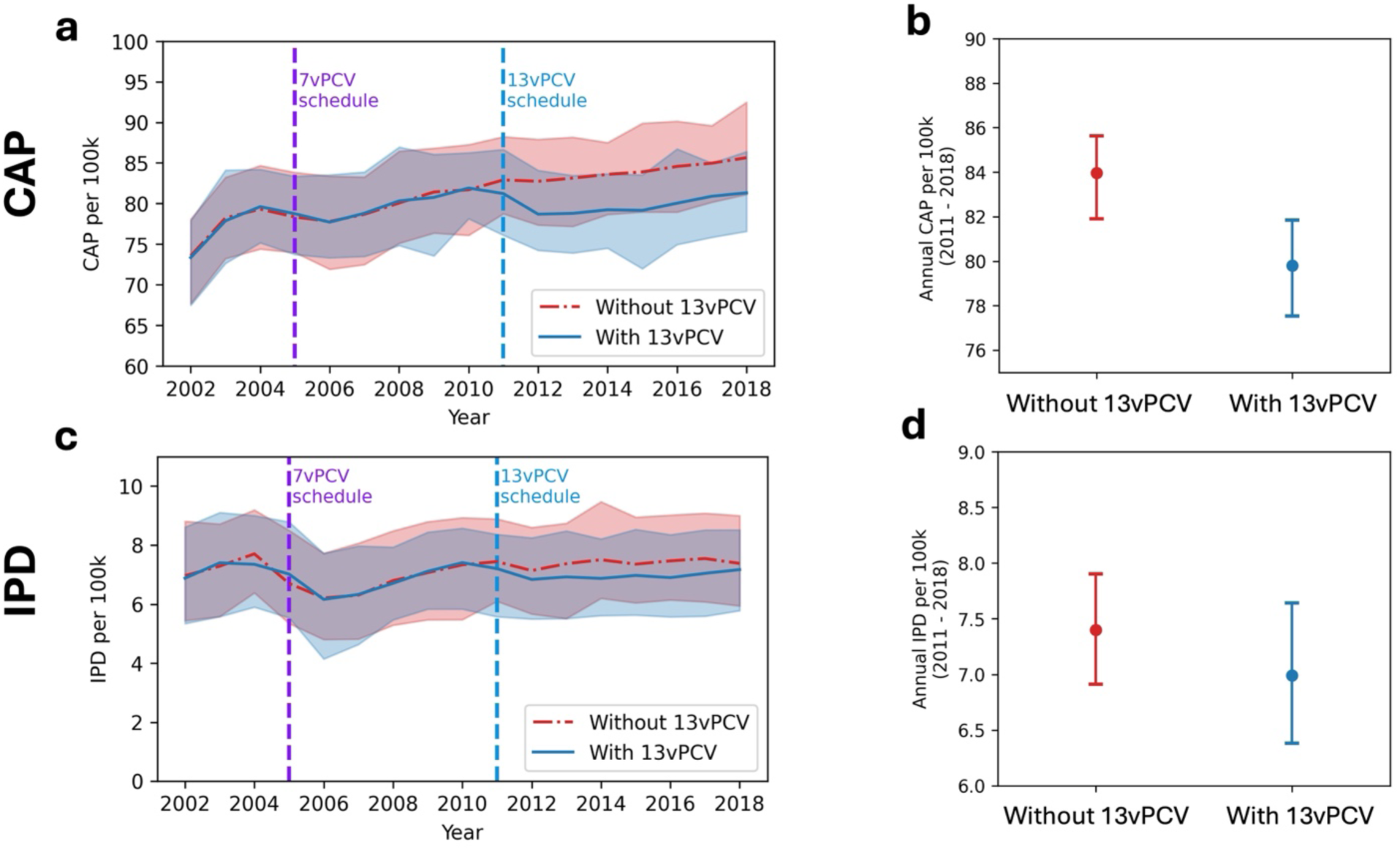
Annual IPD and CAP incidence, per 100,000 between 2002–2018. Panels a and b show the annual CAP burden per 100,000 population, and panels c and d show the annual IPD burden per 100,000 population. Panels a and c show the annual disease burden per 100,000 population with and without 13vPCV scenarios. Panels b and d show the average annual disease burden per 100,000 population for the period 2011–2018.

## 4. Discussion

This paper describes a novel modelling framework that realistically simulates pneumococcal carriage and disease outcome dynamics for evaluation of vaccine strategies. Its development was prompted by the need for a comprehensive model that realistically captures heterogeneities in serotypes, vaccines, and host characteristics to compare effectiveness of different vaccine schedules and inform public health decisions for a given population. Our framework has addressed these challenges, enabling modelling of individual serotype transmission in large populations without introducing biases arising from clustered representation of serotypes in carriage and disease on the basis of their inclusion in a particular vaccine. Moreover, the model has sufficient flexibility to be calibrated to diverse populations and to evaluate multiple vaccine formulations and administration scenarios.

Numerous studies have modelled the impact of pneumococcal conjugate vaccines, with many utilizing a compartmental modelling approach [19–21,29]. While these models account for the multi-valent nature of *Streptococcus pneumoniae* to some degree, they often depend on simplifying assumptions about serotype and host representation [32]. Earlier models tend to use only two serotype groups in relation to the vaccine formulation(s) under study, vaccine-type (VT) and non-vaccine-type (NVT) [19,29]. Recent compartment modelling approaches include more serotype grouping (i.e., seven groupings in Horn et al, 2023 [28]). These compartmental models typically assume that an individual can be co-infected by one VT and one NVT serotype, but not by two VT or two NVT serotypes. This assumption has the potential to alter transmission rates once VT serotype circulation decreases following vaccine introduction. Additionally, these models often assume a homogeneous vaccine-induced immune response to vaccine-type serotypes [19,21,29]. To account for some variations in immune response, Choi et al. (2019) place one vaccine-type serotype with lower vaccine-induced immunity (ST3) into the NVT compartment, assuming all-or-nothing immune response to vaccine-type serotypes [21]. In another compartmental modelling work, Choi et al (2024) include only one new vaccine-type compartment to evaluate multiple vaccines, which requires recalibrating their model [41]. Ojal et al (2017) categorize individuals into broad age groups, which makes it challenging to estimate the impact of vaccine rollout in different age groups [42].

The existing individual-based models tend to model a larger number of serotype groups than the compartmental models (ranging from 15–27 [23–26]). However, it is important to note that these are generic serotypes, not necessarily reflecting the characteristics of actual serotypes, making their application to decision making about public health interventions challenging. In another individual-based model, Van Effelterre et al. (2010) focus solely on carriage in infants under 2 years old [27]. Models that do not cover all age groups cannot estimate the combined impact of multiple vaccines, such as those targeting both paediatric and elderly populations.

Using the modelling framework, we captured a realistic impact of vaccine-induced immunity on carriage and disease outcomes. Our retrospective analysis of pneumococcal disease trends estimated that the 13vPCV paediatric vaccine schedule in the non-Indigenous Australian population reduced the burdens of CAP and IPD by 5% and 5.5%, respectively. This analysis included relevant local context, including host and pathogen-specific heterogeneities and the adult 23vPPV vaccine programme. The model can be applied to consider likely relative benefits of future pneumococcal vaccine schedules for specific populations without introducing simplifying assumptions [43]. Initial PCV developments added new serotypes to those in previous vaccines, enabling the “13vPCV not 7vPCV” style categorisation. However, new vaccines such as 21vPCV do not do this, removing some of the serotypes included in previous vaccines whilst adding new serotypes. By modelling individual serotypes, a key advantage of our approach is that the model can readily incorporate vaccines that do not include all of the serotypes in existing vaccines.

Our modelling approach also comes with limitations. While incorporating pathogen, host, and vaccine-specific heterogeneities can better capture the realistic dynamics of carriage and disease progression, our model requires more data than existing modelling approaches. Due to the lack of historical carriage data in Australia, we made simplifying assumptions about the relationship of infection to disease over time and the relative impact of vaccine efficacy against acquisition compared to disease progression. These assumptions could be tested in settings where granular carriage and disease data are available. In addition, we assumed no natural immunity following an episode of carriage and limited the number of co-infections to two serotypes. Further sensitivity analysis would be needed to understand the impact of these assumptions.

In conclusion, we propose a reliable modelling framework for evaluating vaccine program effectiveness and informing public health strategies by simulating the dynamics of individual serotype transmission and disease progression. While demanding more extensive data than previous models, our model offers greater flexibility and includes evidence-informed vaccine-induced immunogenicity, making it well-suited for assessing new vaccine products and various vaccine strategies for specific populations.

## Data Availability Statement

The model and documentation of how to run the model and input data can be found in the following public repository: https://github.com/nefeltellioglu/abm_pneu_model.

## Data Availability

All data produced in the present study are available upon reasonable request to the authors

## Acknowledgements

This work was funded by the Australian Government Department of Health and Ageing National Immunisation Division. We acknowledge expert input to model structure and assumptions by members of the Australian Technical Advisory Group on Immunisation and the National Immunisation Division. We are grateful to Sanjay Jayasinghe and colleagues at the National Centre for Immunisation Research and Surveillance of Vaccine Preventable Diseases (NCIRS) for assistance with data access and interpretation. The manuscript uses publicly available data sourced from the National Notifiable Diseases Surveillance System. Aggregated national level data on admissions related to CAP (source: Australian Institute of Health and Welfare) and infant IPD (source: Enhanced IPD notifications data) were provided through NCIRS for use in this program of work, with approval from the Communicable Disease Enhanced Surveillance System (CDESS). These data may only be accessed with permission from the primary source.

## Notes

### Competing Interest Statement

The authors have declared no competing interest.

### Author Declarations

This study was funded by the National Immunisation Division of the Australian Government Department of Health, Disability and Ageing to support policy decision making. The model was parameterised using several national data resources provided through the National Centre for Immunisation Research and Surveillance (NCIRS), with approval from the Communicable Disease Enhanced Surveillance System (CDESS). Aggregate national invasive pneumococcal disease data (by serotype and age group) were sourced from the National Notifiable Diseases Surveillance System. Aggregate national data on community acquired pneumonia (CAP) hospitalisations (by age group) were sourced from the Australian Institute of Health and Welfare National Health and Morbidity Database. Infant IPD data were sourced from Enhanced IPD notifications dataset through NCIRS. These data sources may only be accessed with permission from the primary custodian.

### Summary of Updates

We updated the description of the algorithm in the methods section and further highlighted the novelty of the approach in the results section.

