## Supplementary Material for "A flexible agent-based modelling framework of multi-serotype pneumococcal carriage to evaluate vaccine strategies in large populations"

### 1. Immunity model

#### 1.1. Estimation of initial antibody levels

##### 1.1.1. Rapid review of antibody levels

A rapid review was used to search ClinicalTrials.gov for reports published through August 14, 2023. A combination of the following three search terms in "all fields" was used: (1) pneumococcal OR pneumococcus OR "streptococcus pneumoniae" OR "streptococcal pneumoniae" OR "streptococcus p*" OR "streptococcal p*"), (2) vaccine OR vaccines OR vaccination OR immunisation OR immunization OR immunise OR inoculate, (3) conjugate OR valent. Additional relevant studies were retrieved manually from the reference lists of identified articles. One reviewer (Xinghui Chen) screened and determined the inclusion of identified articles. The full texts of potentially eligible studies were evaluated for definitive inclusion.

Eligible studies met the following inclusion criteria: (1) randomized controlled trials (RCTs) investigating any of the five licensed pneumococcal conjugate vaccines (7vPCV, 10vPCV, 13vPCV,15v PCV, 20vPCV) and licensed pneumococcal polysaccharide vaccine containing 23 serotypes (23vPPV) in healthy populations of any age, and (2) results reported as the antibody response assessed by anti-polysaccharide IgG geometric mean antibody concentration (GMC) in micrograms per milliliter (mg/ml) measured with the WHO reference ELISA or with assays validated and bridged to the WHO reference ELISA (e.g. dLIA platform developed by Pfizer, and electrochemiluminescence (ECL) assays). Pneumococcal conjugate vaccines (PCVs) that were not licensed (i.e., 9vPCV, 11vPCV), articles not reporting original research, observational studies, studies focused on immunocompromised patients, interchangeability studies (i.e., first two doses with 7vPCV, then the last dose with 13vPCV), studies on preterm infants, studies on patients with prenatally acquired HIV, studies with only opsonophagocytic assay (OPA) results, and studies in which vaccines were administered subcutaneously were excluded (Table 1).

**Table 1: Studies used in the estimation of the initial antibody levels.**

| Study | First Author & Year | Vaccine | Setting | | | Outcome (IgG GMCs) | |
|---|---|---|---|---|---|---|---|
| | | | Population | Country/region | Schedule | Time points | Assay |
| Infant-toddler series | | | | | | | |
| 1 | Rennels 1998 (*1*) | 7VPCV | Infants | United States | 3 + 1 (2-4-6-12 months) | Before vaccine, post dose 2, post dose 3, pre-booster, post-booster | Standardized ELISA |
| 2 | Shinefield 1999 (*2*) | 7VPCV | Infants | United States | 3 + 1 (2-4-6-12 months) | Before vaccine, post dose 3, pre-booster, post-booster | Standardized ELISA |
| 3 | Black 2000 (*3*) | 7VPCV | Infants | United States | 3+1 (2-4-6-12 months) | Post dose 3 | Standardized ELISA |
| 4 | Eskola 2001 (*4*), Ekstrom 2005 (*5*) | 7VPCV | Infants | Finland | 3+1 (2-4-6-12 months) | 2, 4, 6, 7, 12, 13, 24 months | Kayhty ELISA |
| 5 | Pichichero 2007(*6*) | 7VPCV | Infants | United States | 3 + 0 (2-4-6 months) | Post dose 3 | Standardized ELISA |
| 6 | Scheifele 2006 (*6*) | 7VPCV | Infants | Canada | 3 + 1 (2-4-6-12 months) | Before vaccine, post dose 3 | Standardized ELISA |
| 7 | Kim 2007 (*7*) | 7VPCV | Infants | South Korea | 3 + 1 (2-4-6-12 months) | Before vaccine, post dose 2, post dose 3 | Standardized ELISA |
| 8 | Tapiéro 2013 (*8*) | 7VPCV | Infants | Canada | 3 + 1 (2-4-6-12 months) | Post dose 3, pre-booster, post-booster | Standardized ELISA |
| 9 | Miller 2011 (*9*) | 7VPCV | Infants | United Kingdom | 2 + 1 (2-4-13 months) | Post-booster | Standardized ELISA |
| 10 | Bryant 2010 (*10*) | 7VPCV & 13vPCV | Infants | United States | 3 + 1 (2-4-6-12 months) | Post dose 3, pre-booster, post-booster | Standardized ELISA |

| Study | First Author & Year | Vaccine | Setting | | | | Outcome (IgG GMCs) | |
|---|---|---|---|---|---|---|---|---|
| | | | Population | Country/region | Schedule | | Time points | Assay |
| 11 | Huang 2012 (11) | 7VPCV & 13vPCV | Infants | Taiwan | 3 + 1 (2-4-6-15 months) | | Post infant series, post-booster | Standardized ELISA |
| 12 | Snape 2010 (12) Rodgers 2013 (13) | 7VPCV & 13vPCV | Infants | United Kingdom | 2 + 1 (2-4-12 months) | | Post dose 2, pre-booster, post-booster | Standardized ELISA |
| 13 | Dagan 2013 (14) | 7VPCV & 13vPCV | Infants | Israel | 3 + 1 (2-4-6-12 months) | | Post dose 3, post-booster | Standardized ELISA |
| 14 | Weckx 2012 (15) | 7VPCV & 13vPCV | Infants | Brazil | 3 + 1 (2-4-6-12 months) | | Post dose 3, post-booster | Standardized ELISA |
| 15 | Zhu 2016 (16) | 13vPCV | Infants | China | 3 + 1 (2-4-6-12 months) | | Post dose 3, post-booster | Standardized ELISA |
| 16 | Yeh 2010 (17) | 7VPCV & 13vPCV | Infants | United States | 3 + 1 (2-4-6-12 months) | | Post dose 3, pre-booster, post-booster | Standardized ELISA |
| 17 | Payton 2013 (18) | 7VPCV & 13vPCV | Infants | United States | 3 + 1 (2-4-6-12 months) | | Post dose 3, post-booster | Standardized ELISA |
| 18 | Kim 2013 (19) | 7VPCV & 13vPCV | Infants | South Korea | 3 + 1 (2-4-6-12 months) | | Post dose 3, post-booster | Standardized ELISA |
| 19 | Rodgers 2013 (13) Diez-Domingo 2013 (20) | 7VPCV & 13vPCV | Infants | Spain | 3 + 1 (2-4-6-15 months) | | Post dose 2, post dose 3, post-booster | Standardized ELISA |
| 20 | Vanderkooi 2012 (21) | 13vPCV | Infants | Canada | 3 + 1 (2-4-6-12 months) | | Post dose 3, post-booster | Standardized ELISA |
| 21 | Martinón-Torres 2012 (22) | 13vPCV | Infants | Spain | 3 + 1 (2-4-6-15 months) | | Post dose 2, post dose 3, post-booster | Standardized ELISA |
| 22 | Rodgers 2013 (13) | 13vPCV | Infants | Mexico | 3 + 1 (2-4-6-12 months) | | Post dose 2, post dose 3, post-booster | Standardized ELISA |
| 23 | Temple 2019 (23) | 13vPCV | Infants | Vietnam | 2 + 1 (2-4-9 months) | | Post dose 2, post-booster | Standardized ELISA |

| Study | First Author & Year | Vaccine | Setting | | | Outcome (IgG GMCs) | |
|---|---|---|---|---|---|---|---|
| | | | Population | Country/region | Schedule | Time points | Assay |
| 24 | Gutiérrez 2013 (*24*) | 13vPCV | Infants | Mexico | 3 + 1 (2-4-6-12 months) | Post dose 2, post dose 3, post-booster | Standardized ELISA |
| 25 | Goldblatt 2018 (*25*) | 13vPCV | Infants | United Kingdom | 2 + 1 (2-4-12 months) or 1 + 1 (3-12 months, not included) | Post dose 2, post-booster | Standardized ELISA |
| 26 | Greenberg 2018 (*26*) | 13vPCV & 15VPCV | Infants | United States, Canada, Finland, Israel, and Spain | 3 + 1 (2-4-6-12 months) | Post dose 3, pre-booster, post-booster | Pn ECL* |
| 27 | Korbal 2024 (*27*) | 13vPCV & 20VPCV | Infants | Australia, Czechia, Denmark, Estonia, Finland, Italy, Netherlands, Norway, Poland, Russian Federation, Slovakia | 2 + 1 (2-4-12 months) | Post dose 2 | dLIA# |
| 28 | Senders 2021 (*28*) | 13vPCV & 20VPCV | Infants | United States | 3 + 1 (2-4-6-12 months) | Post dose 3, pre-booster, post-booster | dLIA# |
| Elderly | | | | | | | |
| 1 | Kawakami 2016 (*29*) | 23VPPV | 65+ | Japan | 1 dose | Before vaccination, post vaccination | Pn ECL* |
| 2 | Ermlich 2018 (*30*) | 23VPPV, 13vPCV, or 15VPCV | 65-74 yrs | Canada, Denmark, Israel, Norway, Poland, Spain, Sweden and United States | 1 dose | Before vaccination, post vaccination | Pn ECL* |
| 3 | Ermlich 2018 (*30*) | 23VPPV, 13vPCV, or 15VPCV | 75+ | Canada, Denmark, Israel, Norway, Poland, Spain, Sweden and United States | 1 dose | Before vaccination, post vaccination | Pn ECL* |

* Pn ECL: Pneumococcal electrochemiluminescence assay

### dLIA: Luminex-based direct immunoassay platform

Geometric mean antibody concentration (GMC) with confidence interval was extracted for immunoglobulin G (IgG) antibody for all eligible studies. We captured heterogeneity of IgG values between studies by categorizing the magnitude of antibody responses into three groups (low, moderate and high) for each serotype, vaccine product and dose combination across multiple studies (Tables 2 and 3). The low response group fell into the bottom quartile, the high response into the top quartile and the moderate response into the two middle quartiles. The final group designation of a serotype was then based on their most common presence within a group. As vaccines induce lower protection against serotype 3 (ST3), we introduced an ST3 group in addition to low, moderate and high groups.

From the immunity model, we assigned initial mean antibody levels for each vaccine type after each dose. Antibody levels after the first dose for children for any vaccine type were unavailable in the literature. Therefore, we extrapolated antibody levels after the first dose using the antibody levels after the second and third doses (Fig 1). In the extrapolation, we estimated post-dose 1 antibody levels by multiplying post-dose 2 mean antibody levels by the fold-decrease from post-dose 3 to pose-dose 2. We also assumed log variance as 0.4 for every category in post-dose 1 antibody levels by 7vPCV and 13vPCV.

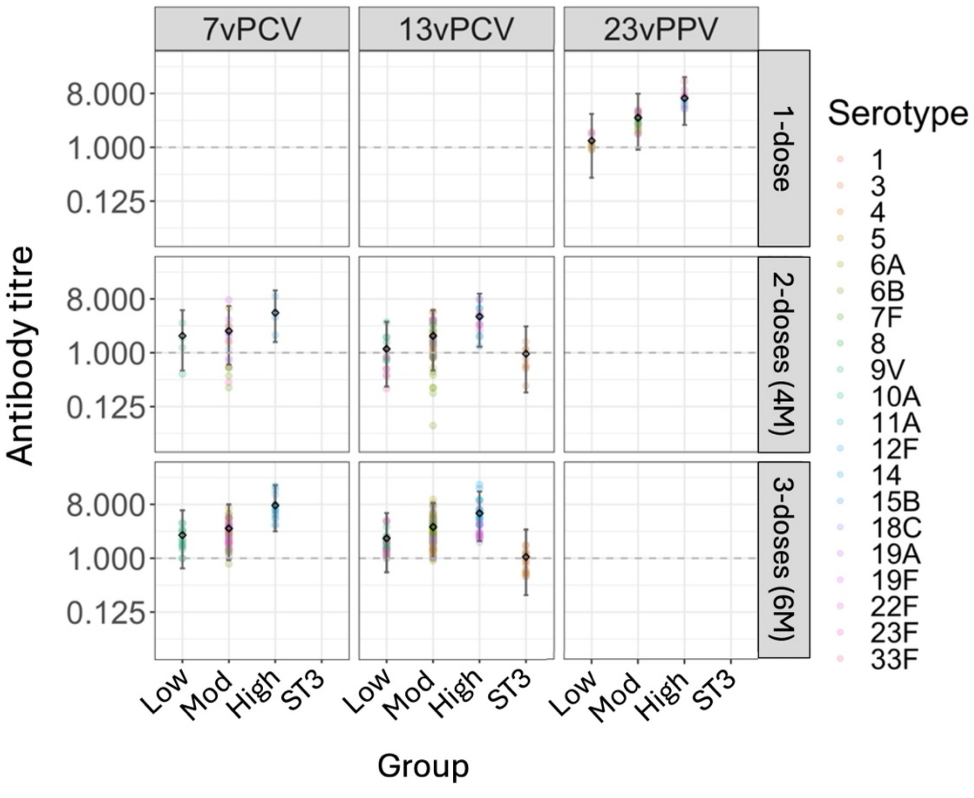

**Fig 1. Antibody distributions**. Mean and confidence intervals from literature for antibody responses by serotype (colour) are shown for each vaccine product, schedule, and group. Lognormal distributions were specified for each combination, such that variability across the various studies was reasonably captured. Black error bars show 2.5–97.5 percentiles for each distribution.

**Table 2. Log antibody distributions**. Mean and variance from literature for antibody responses are shown for each vaccine product, schedule, and group. Lognormal distributions are assumed for each combination, so variability across the various studies was reasonably captured.

| Vaccine | Schedule | Group | Log_Mean | Log_Variance |
|---|---|---|---|---|
| **7vPCV** | Post 1 dose (2 months)* | Low | -0.012313 | 0.400000 |
| **7vPCV** | Post 1 dose (2 months)* | Mod | 0.039943 | 0.400000 |
| **7vPCV** | Post 1 dose (2 months)* | High | 0.272995 | 0.400000 |
| **7vPCV** | Post 2 doses (4 months) | Low | 0.479400 | 0.352060 |
| **7vPCV** | Post 2 doses (4 months) | Mod | 0.676599 | 0.332340 |
| **7vPCV** | Post 2 doses (4 months) | High | 1.410965 | 0.258904 |
| **7vPCV** | Post 3 doses (6 months) | Low | 0.726636 | 0.327336 |
| **7vPCV** | Post 3 doses (6 months) | Mod | 1.000708 | 0.299929 |
| **7vPCV** | Post 3 doses (6 months) | High | 1.932818 | 0.206718 |
| **13vPCV** | Post 1 dose (2 months)* | Low | -0.419364 | 0.400000 |
| **13vPCV** | Post 1 dose (2 months)* | Mod | -0.158107 | 0.400000 |
| **13vPCV** | Post 1 dose (2 months)* | High | 0.279324 | 0.400000 |
| **13vPCV** | Post 1 dose (2 months)* | ST3 | -0.264715 | 0.400000 |
| **13vPCV** | Post 2 doses (4 months) | Low | -0.052441 | 0.405244 |
| **13vPCV** | Post 2 doses (4 months) | Mod | 0.477809 | 0.352219 |
| **13vPCV** | Post 2 doses (4 months) | High | 1.260359 | 0.273964 |
| **13vPCV** | Post 2 doses (4 months) | ST3 | -0.253866 | 0.425387 |
| **13vPCV** | Post 3 doses (6 months) | Low | 0.600220 | 0.339978 |
| **13vPCV** | Post 3 doses (6 months) | Mod | 1.059157 | 0.294084 |
| **13vPCV** | Post 3 doses (6 months) | High | 1.617034 | 0.238297 |
| **13vPCV** | Post 3 doses (6 months) | ST3 | -0.158719 | 0.415872 |
| **23vPPV** | Post 1 dose | Low | -0.020226 | 0.402023 |
| **23vPPV** | Post 1 dose | Mod | 1.143825 | 0.285618 |
| **23vPPV** | Post 1 dose | High | 1.703034 | 0.229697 |
| **23vPPV** | Post 1 dose | ST3 | -0.590378 | 0.459038 |

*The post-dose 1 antibody levels by 7vPCV and 13vPCV vaccines were extrapolated from post-dose 2 and post-dose 3 antibody levels post-7vPCV and 13vPCV vaccination due to the missing data in the literature.

**Table 3. Serotypes in each group.** Serotypes are classified into low, moderate (mod), high, and ST3.

| Vaccine | Group | Serotypes |
|---|---|---|
| **7vPCV** | Low | 9V |
| **7vPCV** | Mod | 4, 6B, 18C, 19F, 23F |
| **7vPCV** | High | 14 |
| **13vPCV** | Low | 9V, 23F |
| **13vPCV** | Mod | 4, 6B, 18C, 1, 5, 6A, 7F, 19A |
| **13vPCV** | High | 14, 19F |

| | | |
|---|---|---|
| **13vPCV** | ST3 | 3 |
| **23vPPV** | Low | 4, |
| **23vPPV** | Mod | 6B, 9V, 19F, 23F, 1, 5, 7F, 2, 11A, 22F, 17F, 33F, 8, 12F, 10A, 15B, 20, 9N* |
| **23vPPV** | High | 14, 18C, 19A |
| **23vPPV** | ST3 | 3 |

*Serotypes 2, 11A, 22F, 17F, 33F, 8, 12F, 10A, 15B, 20, 9N were grouped into moderate category due to the missing information in the literature.

#### 1.2. Waning rates

In the literature reported in Table 1, we looked at the half-lives by serotype across different vaccine product, dosing schedules, and age for studies with multiple post-vaccination timepoints (Fig 2). We calculated the mean decay rate and mean half-life across vaccine products and doses. Based on this analysis, the half-lives of all products were set as 600 days for adults, 125 days for children.

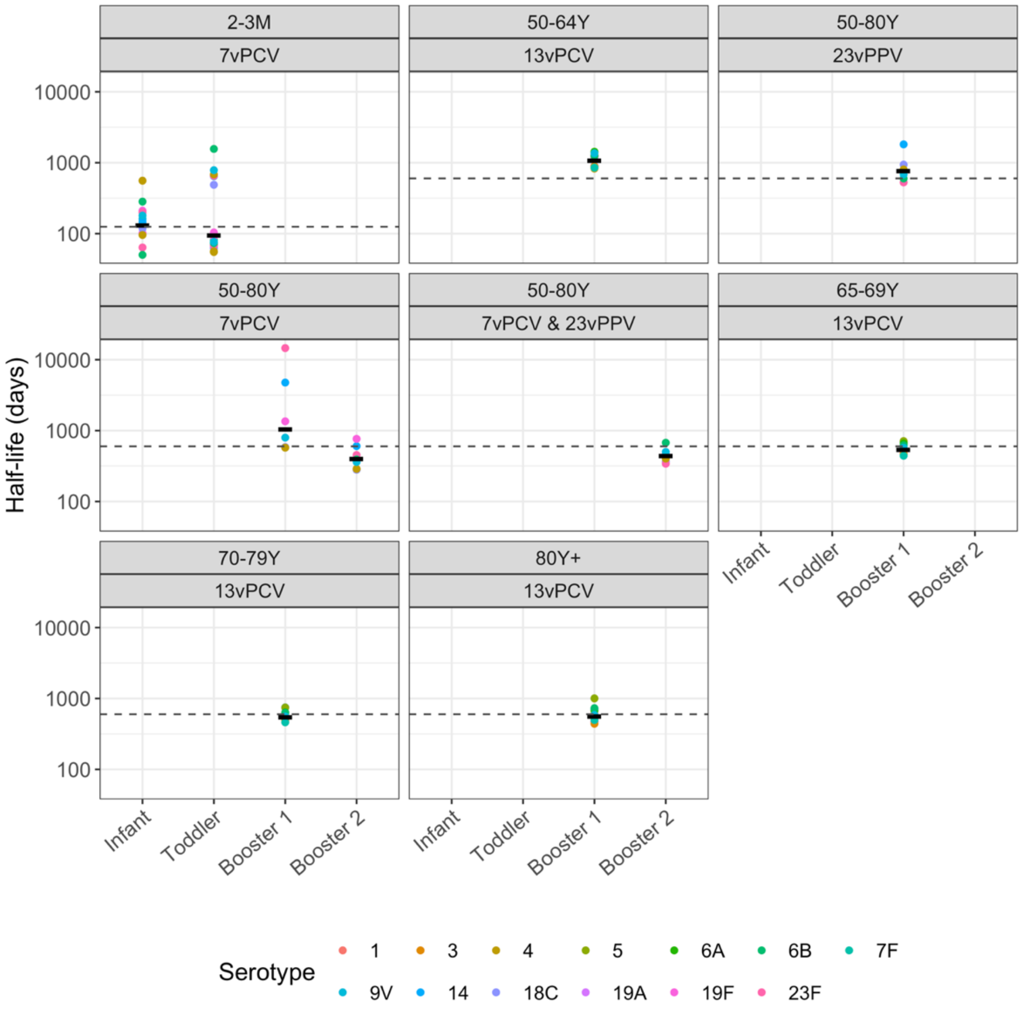

**Fig 2: Antibody half-life assumptions**. Data show estimated half-lives from literature by serotype across different vaccine product, dosing schedules, and age. Black dashes

show mean for each group, and dashed lines show the assumed antibody half-lives used in subsequent modelling (600 days for adults, 125 days for children).

### 2. Transmission model

We constructed a transmission model that incorporates age-related population dynamics. The fundamental model structure draws inspiration from the modelling framework developed by Geard, et al. (2015) (*31*). The model only considers age structure meaning that we do not model household transmission dynamics. Within this model, we create a synthetic population using specified demographic parameters, and then simulate disease transmission within this synthetic population. Through this framework, we perform carriage transmission simulations spanning an extended time horizon, such as decades, while considering realistic population dynamics including population growth and migration.

#### 2.1. Population model

The population model requires the following inputs: initial population size, initial age distribution data, age-specific death rates, annual net birth rates, annual migration rates, and age distribution in the emigrated population (Table 4).

**Table 4: Population model parameters**

| Parameter | Description |
|---|---|
| **Population size** | Number of individuals in the population at the start of the simulation. |
| **Birth rate** | Annual rate of change in population size due to natural increase. |
| **Migration rate** | Annual rate of change in population size due to immigration. |
| **Initial age distribution** | Age distribution at the start of the simulation. |
| **Age-specific mortality probabilities** | Annual probabilities of death given age of an individual. |
| **Age-distribution of the migrated population** | Age-distribution of the emigrated population. |

#### 2.2. Carriage transmission

Carriage of serotypes is determined in multiple steps. We first determine if an individual will be exposed to any serotype, we next determine the serotype, then assess the individual’s serotype-specific vaccine-induced antibody levels and current infections, to determine if the individual gets infected by a serotype.

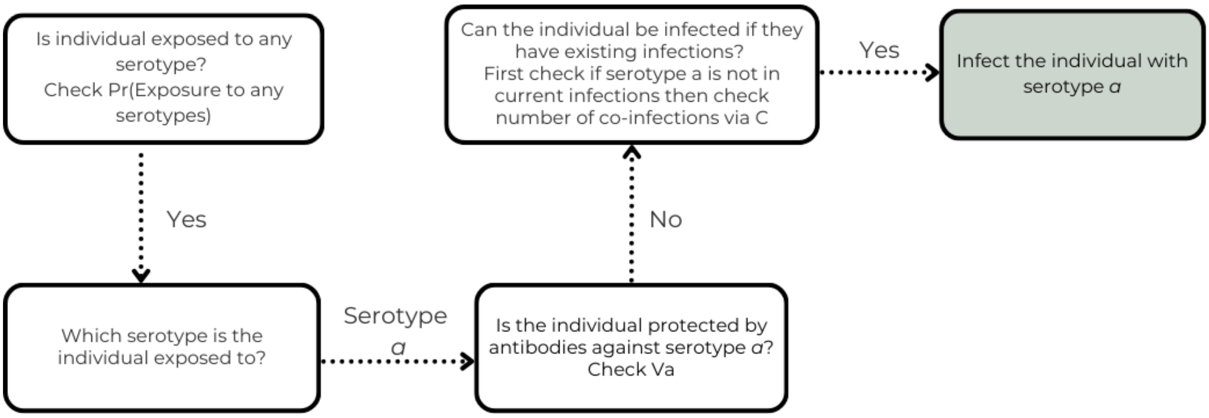

**Fig 3: Flowchart describing the process of an individual acquiring a serotype in a single time step.**

##### *2.2.1. Serotype representation*

In our model, we consider 97 serotypes: serotypes 1, 2, 3, 4, 5, 6A, 6B, 6C, 6D, 6H, 7F, 7A, 7B, 7C, 7D, 8, 9A, 9L, 9N, 9V, 10F, 10A, 10B, 10C, 10D, 11A, 11B, 11C, 11D, 11E, 12F, 12B, 13, 14, 15F, 15A, 15B, 15C, 16F, 16A, 17F, 17A, 18F, 18A, 18B, 18C, 19F, 19A, 19B, 19C, 20A, 20B, 21, 22F, 22A, 23F, 23A, 23B, 24F, 24A, 24B, 24C, 25F, 25A, 27, 28F, 28A, 29, 31, 32F, 32A, 33F, 33A, 33B, 33C, 33D, 34, 35F, 35A, 35B, 35C, 35D, 36, 37, 38, 39, 40, 41F, 41A, 42, 43, 44, 45, 46, 47F, 47A, 48. In our model, serotype characteristics vary in terms of transmission rates and acquired antibody levels following vaccination. Characteristics (e.g. duration of carriage) of non-vaccine serotypes are the same, however, including individual serotypes in the model eliminates any bias of co-infection which is explained further in the next section.

##### *2.2.2. Co-infection of a host by multiple serotypes*

In previous modelling studies, the number of serotypes that can co-infect individuals varies. Many previous studies assume that individuals can carry a maximum of two serotypes at a given time (*32–34*). They assume that only serotypes belonging to different categories such as vaccine type and non-vaccine type, can co-infect a host. For example, Choi et al. (2019) assume that individuals can be infected by three serotypes, as long as the serotypes are from different categories (vaccine type 1, vaccine type 2, or non-vaccine type) (*35*). However, this assumption is a model simplification rather than being based on biological evidence of within-host competition.

Allowing co-infection between categories while prohibiting co-infection within a category can introduce a bias in transmission. As a hypothetical scenario, we can imagine that when the vaccine-type serotypes are eliminated or reduced significantly from a population after an effective vaccination rollout, non-vaccine-type serotypes dominate the carriage in the population. However, due to the modelling assumption of no co-infection among non-vaccine type serotypes, the transmission of serotypes becomes lower compared to the transmission before the vaccination rollout. Therefore, this assumption may artificially conclude a lower level of carriage after the vaccination rollout. In order to eliminate this bias, we consider every individual serotype in our model. We include a parameter describing the maximum number of co-infections in our model.

Many of the previous models assume a reduction in susceptibility to future infections when an individual is already infected by a serotype (32, 33, 35, 36). Nurhonen et al. (2013) assume a 74% or 80% reduction depending on the serotype groups while Choi et al. (2019) assume a reduction ranging between 0% to 100% depending on the serotype groups and the age of the individuals. In our model, we introduce a reduction in susceptibility parameter (Table 1, main text).

###### *2.2.3. Vaccination*

Our framework accommodates historical vaccination schedules, formulations, and uptake. It takes inputs of antibody response to vaccination from the immunological model and incorporates waning of vaccine-induced immunity over time. It uses antibody levels at the time of an individual's exposure to determine the likelihood of infection acquisition. Model outputs reporting newly-acquired infections provide inputs for the clinical model.

The model can simulate any age-specific vaccine rollout with a given number of doses and individual's day of vaccination. When an individual is vaccinated, their vaccine status is updated with vaccine name, dose, and final vaccination time to be later used in the acquisition and developing disease events.

In our model, we assign a random quantile value for every individual that remains constant for individuals throughout the simulation. When an individual is exposed to a vaccine-type serotype after vaccination, we first calculate their "initial" antibody levels (antibody levels that they would have to that specific serotype just after the last dose of vaccination). We calculate their "initial" antibody levels by pinpointing their quantile value in a lognormal distribution with the mean and standard deviation of the log antibody levels of the serotype category that serotype belongs to. Then we calculate effective antibody levels at the time by multipling "initial" antibody levels by the waning rate, accounting for the time since the last dose of vaccination. Using the effective antibody level, we calculate the corresponding probability of acquisition or developing disease (Fig 4).

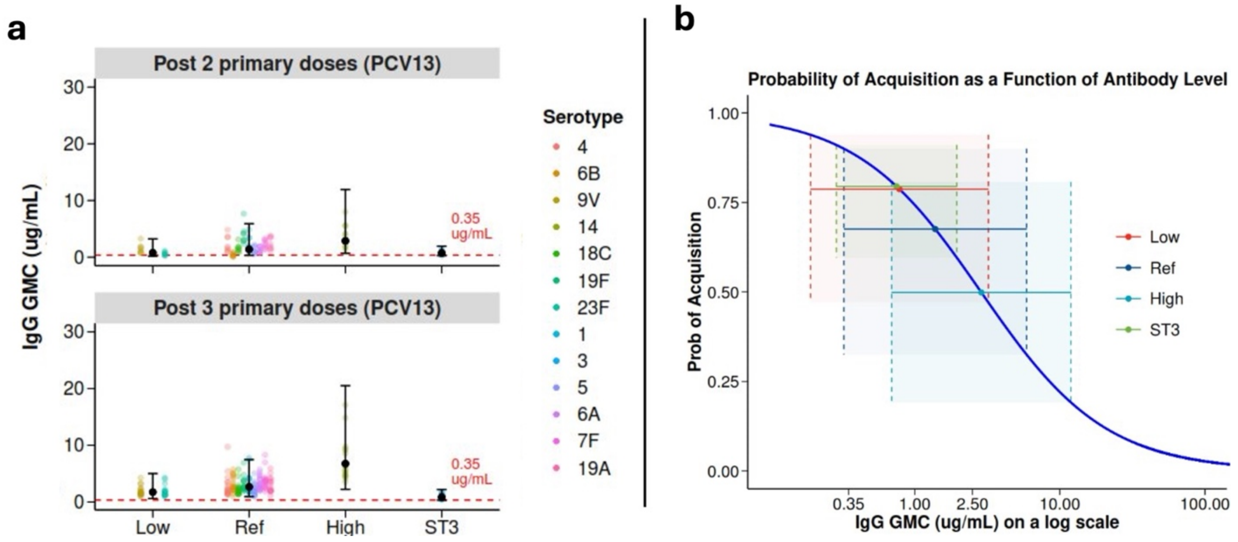

**Fig 4. Initial antibody levels post 2 and 3 doses of 13vPCV vaccine (a) and an exemplar probability of acquisition given antibody levels (b).**
In the vaccine rollout, we assumed that individuals receiving the first dose on time would also receive the following doses on time. For the years that vaccine types are changed, i.e., 7vPCV to 13vPCV in 2011, we assumed that individuals who already received at least one dose of the earlier vaccine type, received the following doses from the same vaccine type.

### 3. Clinical model

In our model, at every time step, we record individuals who develop hospitalized community acquired pneumonia (CAP) or invasive pneumococcal disease (IPD). We assume that individuals can develop disease at any time during their infectious period, therefore, we check every infected individual in every time step. We allow only one disease outcome for every infectious period. When an individual develops a disease based on the probability of developing a disease, we record the disease outcome as either hospitalized CAP or IPD using the age-specific IPD ratio among IPD or CAP cases input parameter (Fig 5).

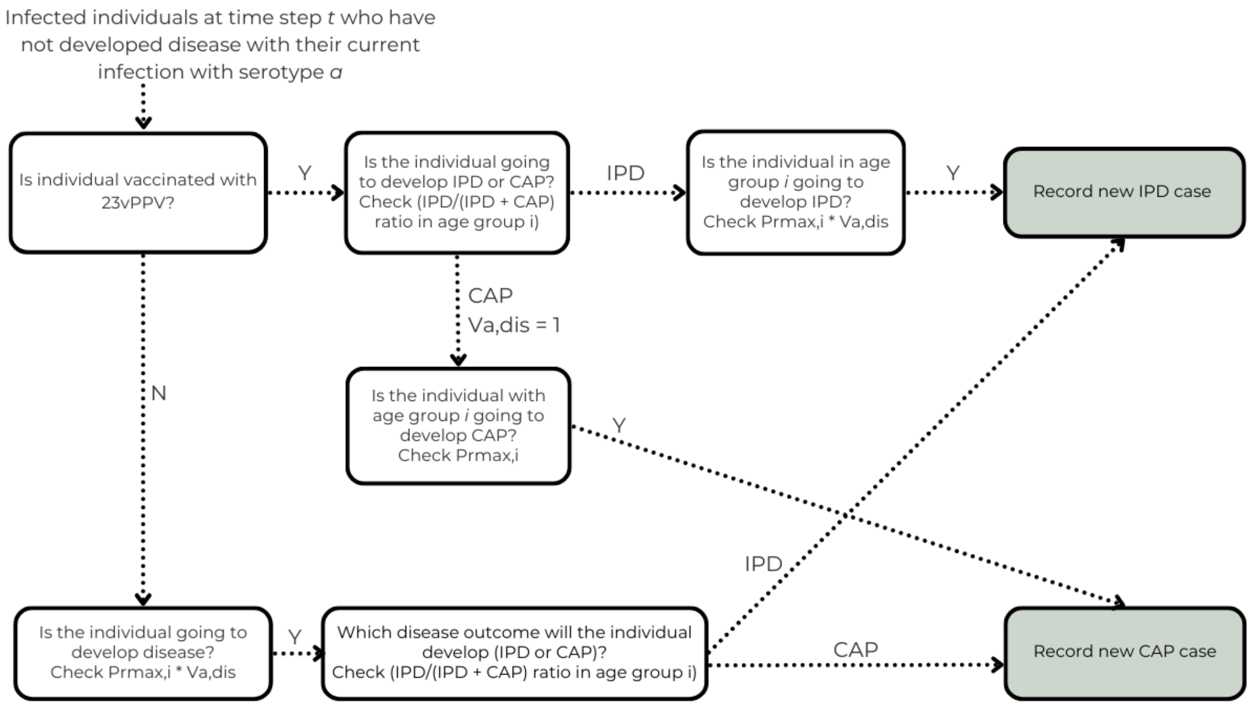

**Fig 5: Flowchart describing the process of an individual developing disease from their current infection.** For individuals vaccinated with 23vPPV, vaccine-induced antibodies only provide protection against IPD. Y = Yes, N = No.

### 4. Case study

#### 4.1. Population model

Initially, we calibrated the demographic structure in our model to align with the demographics of the non-Indigenous Australian population. We then compared the age-related characteristics observed in our synthetic population with data from

Australia. Subsequently, we conducted simulations over a 20-year period, spanning from 2002 to 2022, with an initial population size of 1,000,000 (Table 5).

**Table 5: Population model parameters in the case study**

| Parameter | Description | Values/ References |
|---|---|---|
| **Population size** | Number of individuals in the population at the start of the simulation. | 1,000,000 |
| **Birth rate** | Annual rate of change in population size due to natural increase. | 2002–2042 actual and projected data taken from www.macrotrends.com |
| **Migration rate** | Annual rate of change in population size due to immigration. | 2002–2042 actual and projected data taken from www.macrotrends.com |
| **Initial age distribution** | Age distribution at the start of the simulation. | 2002 data taken from Australian Bureau of Statistics (*37*) |
| **Age-specific mortality probabilities** | Annual probabilities of death given age of an individual. | 2002 data taken from Australian life tables, 2002–2004, Australian Bureau of Statistics (*37*) |
| **Age-distribution of the migrated population** | Age-distribution of the emigrated population. | 2018–2019 data taken from Australian Bureau of Statistics (*37*) |

We initialized the age distribution of the population based on 2002 non-Indigenous Australian data (Fig 6). We then compared simulated population outputs in 2012 and 2022 with observed population growth and age distribution reports from 2012 and 2022 (Figs 7 and 8).

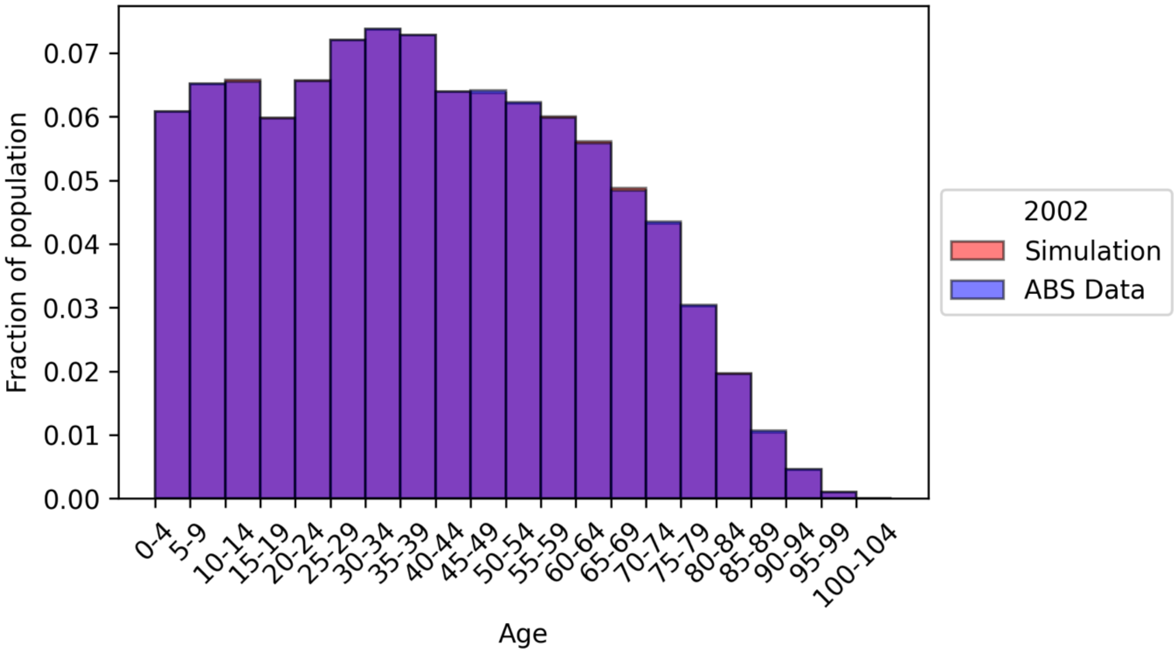

**Fig 6. Initialization of age distribution in 2002.** The age distribution of the model (Simulation, orange) is initialized using the non-Indigenous age distribution data taken from the Australian Bureau of Statistics (ABS Data, blue).

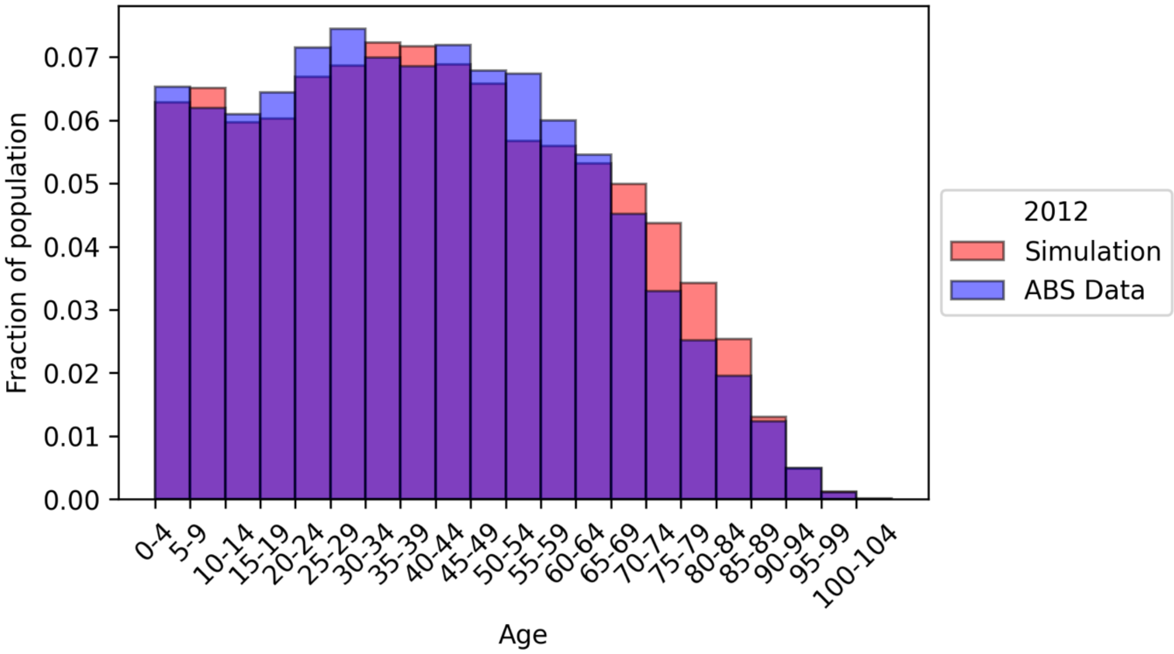

**Fig 7. Comparison of age distributions in 2012.** The age distribution of the model captured in 2012 (Simulation, orange) reflects the 2012 non-Indigenous Australian age structure taken from the Australian Bureau of Statistics (ABS Data, blue).

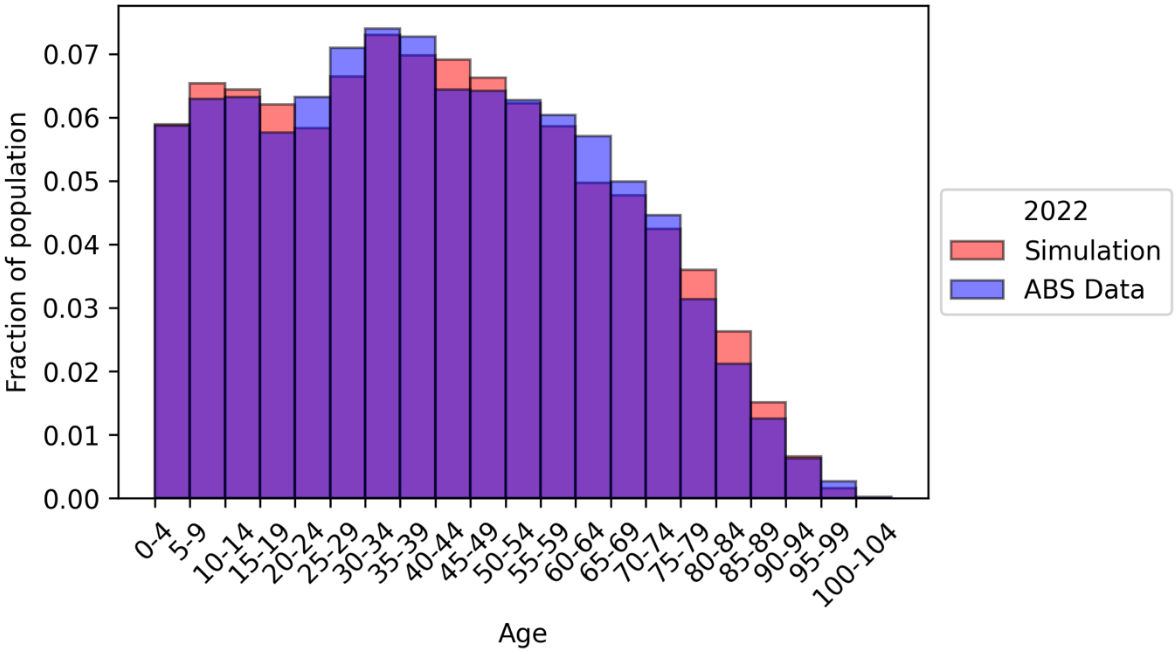

**Fig 8. Comparison of age distributions in 2022.** The age distribution of the model captured in 2022 (Simulation, orange) reflects the 2022 non-Indigenous Australian age structure taken from the Australian Bureau of Statistics (ABS Data, blue).

Growth of the modelled population through birth and migration also reflects the observed Australian non-Indigenous population dynamics. From 2002 to 2012, a 15.3% (16%) increase in size is observed in the simulation (concordant with the actual population). Over the 20-year period from 2002 to 2022, this increase reaches 29.5% (34%), again matching Australian data.

#### 4.2. Carriage transmission

##### *4.2.1. Age-specific duration of infection*

We indirectly account for the accumulation of protection against pneumococcus over the life course by incorporating the observed reduction of carriage duration throughout late childhood and adult life, as shown in Fig 9 (*33*, *35*, *38*, *39*).

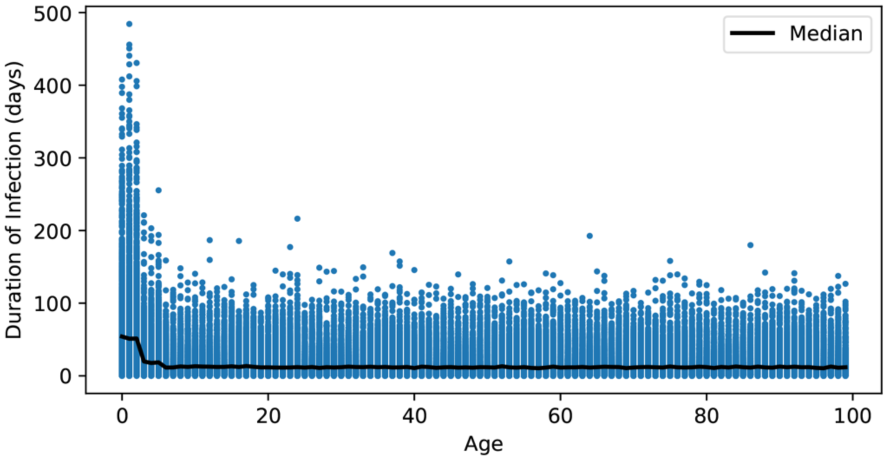

**Fig 9. Age-specific duration of infection.** Duration of infection is modelled as age-specific and values are randomly selected from an exponential distribution with mean values of 72 days for 0–2-year-olds, 28 days for 2–4-year-olds, 18 days for 5–17-year-olds, and 17 days in 18+-year-olds. Sampled durations and median of 100,000 infected individuals from a single population is presented.

##### *4.2.2. Vaccine Coverage*

The model simulates pneumococcal vaccine rollout from the commencement of the nationally funded 7vPCV program for all infants in non-Indigenous Australian population in 2005, with 7vPCV, 13vPCV and 23vPPV (in adults) included. We refer to the data taken from the immunisation coverage annual reports prepared by NCIRS as "NCIRS data" (*40*). We did not include the nationally funded catch-up programs in 2005, when 7vPCV was introduced. While 10vPCV was implemented in some risk groups over this period, uptake was limited and in the interests of parsimony we have not incorporated its use in the model.
Vaccination uptake in children was drawn from Kabir et al. (*41*). From data, we have estimated probabilities of an individual receiving vaccination on time and vaccination occurring after the due date but before the measurement milestone (Figs 10–12). We do not account for the small number of vaccinations received after the milestone.

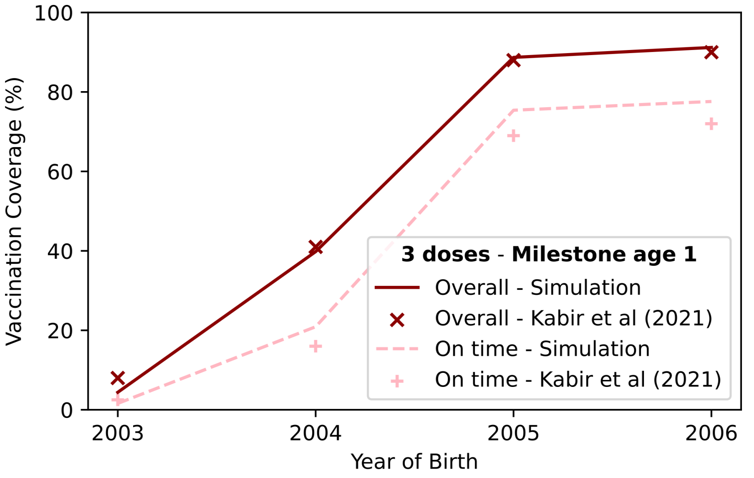

**Fig 10. Comparison of vaccination coverage at 12 months of age between 2003–2006**. For the years 2006 and 2007, on-time vaccination coverage was higher in NCIRS vaccine coverage data than Kabir et al. data which was taken from only two states. Therefore, for the years 2003–2006, we systematically increased the on-time coverage percentages to match the NCIRS vaccine coverage data for the non-Indigenous population (*41*).

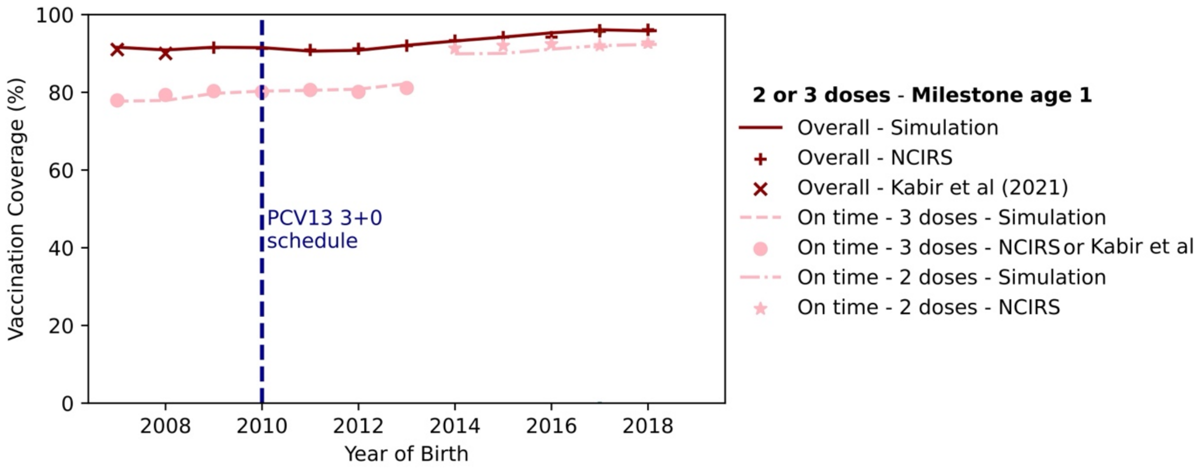

**Fig 11. Comparison of 2 or 3 doses vaccination coverage at 12 months of age between 2007–2019.** Before 2017, overall coverage represents 3 doses coverage in both simulation and data taken from NCIRS Annual Immunisation Coverage Reports. After 2017, overall coverage represents 2 or 3 doses coverage in both simulation and data taken from NCIRS Annual Immunisation Coverage Reports. On-time three doses data between 2007-2008 is taken from Kabir et al, and the on-time three doses data between 2009-2018 is taken from NCIRS. In the NCIRS data, two and three doses coverage data were not provided for the same year. Therefore, in our model, we assumed that individuals who received two doses on time would also receive the third dose on-time. To match the data, we introduced a jump in on-time coverage percentage in 2014.

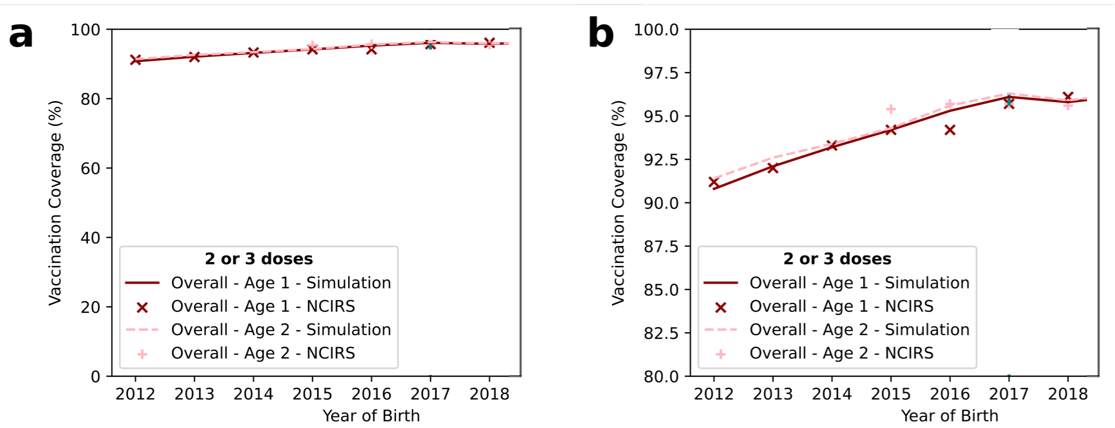

**Fig 12. Comparison of vaccination coverage at 12 and 24 months of age between 2012–2020.** Before 2017, overall coverage represents 3 doses coverage in both simulation and NCIRS data. After 2017, overall coverage represents 2 or 3 doses coverage in both simulation and NCIRS data. Panel b represents a close-up of the data in Panel a.

PCV indications for adults under 70 years are restricted to risk groups and so in our model of the total population, we only considered vaccination with 23vPPV. Vaccination uptake of 23vPPV was drawn from Frank et al. (2020), noting that such data are scarce for Australia (*42*). We focused on capturing the data presented in Study 1 of Frank et al. (2020), which was designed to calculate annual pneumococcal vaccination uptake (Fig 13). Our modelled vaccinated status shows some divergence from the `ever vaccinated' individuals of Frank et al. (2020), as our “ever vaccinated” increases over time, as expected with a continuous vaccination program, while theirs decreases, without any explanation of the reason for this observation.

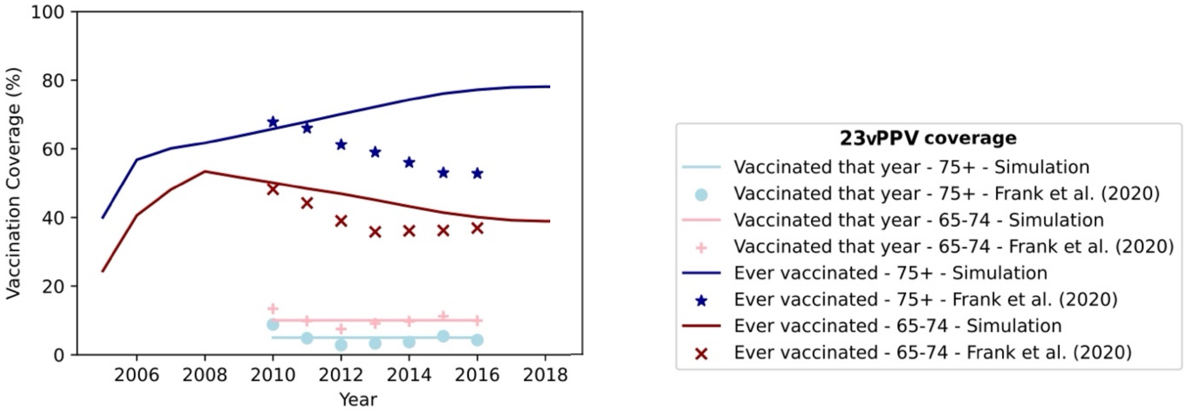

**Fig 13. 23vPPV vaccine coverage.** Data shown for ‘vaccinated that year’ (pink and blue symbols and lines) were used to determine the probability of an individual aged 65 and over being vaccinated.

For the case study, we consider vaccination schedules shown in Table 6. For catch up program, we randomly select 30% of the individuals aged between 1-3 who are already vaccinated by 7vPCV to receive a single dose of 13vPCV. Once individuals receive a single catch-up dose of 13vPCV, we assume their protection is determined solely by the antibody levels generated by this 13vPCV dose, rather than by any antibodies previously induced by 7vPCV.

**Table 6: Vaccine schedules used in simulation study.** The on-time and late coverage fractions are calibrated to the milestone age coverage data from NCIRS, Kabir et al (2021), Frank et al. (2020).

| Vaccine | Years | Schedule | Age range (years) | Annual on-time coverage fraction | Annual late coverage fraction | Previous vaccination |
|---|---|---|---|---|---|---|
| 7vPCV | 2005–2010 | 2, 4 and 6 months of age | 0–2 | 0.109, 0.749, 0.779, 0.779, 0.779, 0.793, 0.803 | 0.221, 0.151, 0.151, 0.151, 0.151, 0.137, 0.127 | — |
| 13vPCV | 2011–2018 | 2, 4 and 6 months of age | 0–2 | 0.801, 0.806, 0.815, 0.893, 0.899, | 0.119, 0.114, 0.119, 0.037, 0.048, | — |

| | | | | | | |
|---|---|---|---|---|---|---|
| | | | | 0.912, 0.916 | 0.046, 0.049 | |
| 13vPCV catchup (single dose) | 2011–2012 | 2 months after individuals' birthday | 1–3 | 0.3 | 0 | 7vPCV |
| 23vPPV | 2005–2018 | 2 months after individuals' birthday | 65+ | 65-74: 0.25, 0.25, 0.2, 0.2, 0.1,..., 0.1<br><br>75+: 0.4, 0.3, 0.1, 0.05,..., 0.05 | 0 | - |
